# Impact of clinical risk factors on white matter microstructure in preterm-born infants: Investigation with diffusion MRI and tractography at term-equivalent age

**DOI:** 10.64898/2026.02.12.26346175

**Authors:** L. Devisscher, Y. Leprince, V. Biran, N. Elbaz, C. Ghozland, P. Adibpour, C. Chiron, S. Neumane, A. Gonzalez Carpinteiro, M. Elmaleh-Bergès, L. Hertz-Pannier, A. Heneau, M. Barbu-Roth, M. Alison, J. Dubois

**Affiliations:** Université Paris-Cité, INSERM, NeuroDiderot, F-75019 Paris, France; Université Paris-Saclay, CEA, NeuroSpin, UNIACT, F-91191 Gif-sur-Yvette, France; Université Paris-Cité, CNRS, Integrative Neuroscience and Cognition Center, F-75005 Paris, France; Assistance Publique-Hôpitaux de Paris - APHP, Robert-Debré Hospital, Neonatal Intensive Care Unit, F-75019 Paris, France; Assistance Publique-Hôpitaux de Paris - APHP, Robert-Debré Hospital, Department of Pediatric Radiology, F-75019 Paris, France

**Keywords:** Prematurity, brain development, maturation and dysmaturation, sensorimotor network, biomarkers, neurodevelopmental disorders, diffusion tensor imaging (DTI)

## Abstract

Premature birth occurs during a phase of intense brain maturation, making white matter (WM) particularly vulnerable to injury. Beyond major lesions, subtle and widespread microstructural alterations also contribute to later neurodevelopmental impairments. We aimed to characterize the impact of key clinical risk factors on global and tract-specific WM microstructure at term-equivalent age (TEA), using 3T-diffusion-MRI data of 111 infants born before 33 weeks of gestation. We implemented a tractography pipeline adapted for use in infants with heterogeneous brain anatomy, including macroscopic abnormalities and lesions, and extracted diffusion tensor imaging (DTI) metrics in sensorimotor tracts: corticospinal tract (CST), superior thalamic radiation (STR), frontal aslant tract (FAT), forceps minor (FMI) and middle cerebellar peduncle (MCP). Associations with risk factors were assessed accounting for age at MRI or global WM microstructure. Tractography succeeded in most infants despite marked anatomical variability and/or overt lesions. Being a male, small for gestational age (SGA) at birth, encountering sepsis and having severe Kidokoro radiological score for WM were associated with altered global WM metrics. At the tract level, CST and STR showed the strongest susceptibility to SGA, prolonged parenteral nutrition, and Kidokoro score. In contrast, associations after adjustment for global WM microstructure suggested that FAT might be less vulnerable than other tracts to extreme prematurity, SGA and invasive ventilation. Findings were partially replicated in infants without macroscopic abnormalities, supporting the presence of WM dysmaturation even in the absence of visible injury. DTI metrics thus provide tract-specific biomarkers of early WM microstructure in preterm infants, which are sensitive to risk factors and could inform targeted prevention and intervention.

## Introduction

Despite recent progress in perinatology, very preterm birth, defined as delivery before 32 weeks of gestational age (GA), remains a major global health concern with a high incidence. It represents a major risk factor for neurodevelopmental disorders, with substantial impact on the long-term quality of life. For instance, in a large French neonatal cohort recruited in 2011, neurodevelopmental outcomes assessed at 5½ years revealed moderate to severe disabilities in 27.8%, 18.7%, and 11.6% of children born at 24–26, 27–31, and 32–34 weeks of gestation respectively (Pierrat et al., 2021).

### Prematurity, neurodevelopmental disorders and brain lesions

Advances in perinatal management over the past decades have led to a marked decline in the incidence of cerebral palsy among children born very preterm. While severe motor impairments can be identified quite early during infancy and toddlerhood, more subtle motor difficulties often emerge later in childhood, particularly when the demands of learning at school increase. These impairments are increasingly recognized as downstream consequences of early disruptions in the progression of critical mechanisms of brain development during the perinatal period (Chung et al., 2020).

Prematurity may lead to characteristic brain lesions resulting from the immaturity and early vulnerability of brain tissues and hemodynamic system, with also a potential role of pro-inflammatory factors altering developmental mechanisms such as myelination (Elbaz et al., 2025). Beside the classical periventricular leukomalacia (PVL) and intraventricular hemorrhage (IVH), which are generally identified by transfontanellar ultrasound in the first days of life, other entities, such as diffuse excessive high signal intensity (DEHSI), reflect more subtle and widespread alterations of white matter (WM) microstructure, which require magnetic resonance imaging (MRI) at term-equivalent age (TEA) to be detected (Inder et al., 2003; Kidokoro et al., 2013).

### Beyond overt brain lesions: the role of dysmaturation

Emerging evidence indicates that even in the absence of overt lesions, preterm infants frequently exhibit neurodevelopmental deficits, particularly in sensorimotor and cognitive functions. This suggests that beyond visible lesions, disturbances in brain maturation processes and progression, often referred to as cerebral dysmaturation, may affect the development of functional brain networks (Volpe, 2009). However, it is still poorly understood what precise mechanisms are at play and contribute to the heterogeneity of outcomes: subtle microstructural alterations, connectivity disruptions, delayed myelination in specific WM tracts or others. Furthermore, the temporal dynamics of dysmaturation, i.e. which pathways are most vulnerable and when, remain largely unexplored. Microstructural lesions may exert dissociated effects on WM maturation of specific pathways and networks, as well as on global WM. And the effects of perinatal risk factors on the modulation of developmental neuroplasticity have been insufficiently studied.

### The promise of advanced neuroimaging

To investigate these questions, one major challenge remains the automatic and reliable analysis of WM pathways in large neonatal cohorts. MRI is the method of choice to explore the preterm developing brain, but data quality and acquisition settings differ substantially in clinical contexts compared with those typically obtained in adults (Dubois et al, 2021). Diffusion MRI offers a non-invasive window into the microstructural integrity of WM (Dubois et al., 2014). In particular, the diffusion tensor imaging (DTI) model allows the measurement of metrics such as fractional anisotropy (FA) and mean diffusivity (MD), which are sensitive markers of brain maturation, capturing complex processes including WM myelination, axonal organization and density. Importantly, even in the absence of overt visible lesions, alterations in these metrics have been consistently reported in preterm infants at TEA compared to full-term neonates, indicating microscopic lesions or microstructural dysmaturation in localized regions (Hüppi et al., 1998). Additional diffusion MRI approaches including tract-based spatial statistics (TBSS), region-of-interest (ROI) analysis, voxel-wise comparison, tractography, and more recently fixel-based analysis, all revealed regions of delayed maturation or disrupted microstructural organization in the preterm brain (e.g., Anjari et al., 2007; Hasegawa et al., 2010; Pannek et al., 2018; Rose et al., 2008; Lepomäki et al., 2013). Moreover, tractography approaches are particularly valuable as they enable the identification of long-distance connections, for example between cerebral lobes, thereby providing an integrative view of the organization and vulnerability of neural networks.

### The impact of prematurity on the sensorimotor network

In the context of prematurity, the cerebral sensorimotor network deserves particular attention. This network comprises both cortical regions (primary somatosensory cortex in the post-central gyrus, primary motor cortex in the pre-central gyrus, secondary somatosensory cortex and premotor cortex, supplementary motor area) and subcortical structures (the thalamus, particularly the ventral lateral and ventral posterior nuclei involved in motor and somatosensory relay respectively; the basal ganglia with caudate nucleus, putamen and globus pallidus; and the cerebellum), which are connected through various WM pathways. It is among the earliest brain systems to mature, with rapid development occurring during the late second and third trimesters of gestation. Consequently, preterm birth occurs during a critical window, when the sensorimotor system is highly vulnerable to disruption. Furthermore, early microstructural abnormalities detected by diffusion MRI in the sensorimotor tracts have been linked to neurodevelopmental impairments observed during infancy and early childhood (Parikh et al., 2021; Neumane et al., 2022), with potential long-term consequences on sensory, motor and cognitive functions. In this study, we then aimed to focus on the main WM tracts essential for sensorimotor function, encompassing both cortico-subcortical and cortico-cortical pathways.

### The impact of perinatal risk factors

Besides, preterm neonates frequently face complex clinical challenges (including respiratory, digestive and infectious complications) which necessitate intensive neonatal care (including respiratory support, nutritional management and treatment of infections). In addition to prematurity itself, a multitude of perinatal risk factors and protective factors may critically influence cerebral maturation and the microstructural development of the brain (Barnett et al., 2018). For instance, higher energy and lipid intake during the first two weeks of life has been associated with greater brain growth and more mature WM microstructure in preterm infants (Schneider et al., 2018), whereas mechanical ventilation can induce oxidative stress through localized cerebral inflammatory response and hemodynamic instability and thereby increase the risk of brain injury (Cannavò et al., 2020). Such interventions, while essential for survival, have been observed to be linked to atypical WM development through inflammatory, metabolic, and hemodynamic mechanisms affecting myelination and neurite organization (Oloomi et al., 2025).

Actually, prolonged invasive ventilation, extended parenteral nutrition, and perinatal sepsis have been associated with lower FA and higher MD in WM on MRI at TEA (Ball et al., 2010; Lee et al., 2014; Shah et al., 2008; Shin et al., 2016). Collectively, these findings support the view that prematurity-related clinical factors contribute to WM microstructural alterations and widespread dysmaturation. Nevertheless, the majority of these studies have either considered ROIs or specific tracts in isolation, leaving open the question of how risk factors may affect entire brain networks. They have largely left unanswered the question of how specific factors differentially impact single tracts within a network.

Sex is also to be considered given the well-documented higher vulnerability of male preterms to moderate-to-severe neurodevelopmental disorders, suggesting a possible protective effect of female sex (Pierrat et al., 2021, Pogribna et al., 2013). Regarding the critical role of GA at birth in shaping brain microstructure, altered WM characteristics have been observed in preterms at TEA compared to full-term neonates (Pannek et al., 2013), and extremely-to-very preterm infants (<32 weeks GA) exhibited less mature microstructural profiles in sensorimotor tracts than moderate-to-late preterms (32–36 weeks GA), suggesting a gradient of WM alterations with GA at birth (Neumane et al., 2022). Besides, higher birth weight appears to exert a protective effect on several brain regions (Pogribna et al., 2013).

### Hypotheses and aims of the study

Contrarily to previous studies comparing preterm and full-term neonates to assess the effect of prematurity, here we aimed to explore whether specific clinical factors might modulate the WM microstructure at TEA in a large cohort of extremely-to-very preterm born infants. The inter-individual variability in DTI metrics was analyzed according to the following variables, selected based on their relevance as established neonatal risk factors (Elbaz et al., 2025): sex, monochorionic twin pregnancy, category of GA at birth, being small for GA at birth (SGA), the occurrence of bronchopulmonary dysplasia (BPD), the use of invasive mechanical ventilation, the presence of necrotizing enterocolitis (NEC), the use of prolonged parenteral nutrition, and perinatal sepsis.

We hypothesized that the first four factors, mainly related to genetics and gestation, might primarily affect the global WM microstructure, while prematurity-related risk factors might rather modulate the differential vulnerability of sensorimotor network tracts given their heterogeneous progression of myelination. Specifically, connections to the frontal lobe may be disproportionately affected because of their delayed maturation, which could render them particularly vulnerable to perinatal complications. Alternatively, their relative immaturity may support later functional reorganization, rather than providing protection against early microstructural injury. Then severe neonatal morbidities were expected to lead to tract-specific alterations in DTI metrics, in relation to the differential maturation progression after preterm birth.

In this study, the precise identification of WM connections further required the development of an automated processing pipeline of diffusion-weighted (DWI) images, tailored to a specific clinical population and including a state-of-the-art tractography method. This allowed us to reliably quantify tract-specific microstructural properties that were related with selected risk factors.

## Materials and Methods

### Cohort

Prematurely born infants admitted immediately after birth to tertiary neonatal intensive care units were enrolled between April 2021 and July 2024 for MRI at Robert-Debré Hospital (Paris, France). Eligibility criteria included preterm birth before 33 weeks of gestation with completion of MRI at TEA as part of the standard procedure [i.e., routine indications: extreme prematurity (<28 weeks gestation) or preterm birth (<37 weeks gestation) accompanied by either abnormal cranial ultrasound findings or clinical concerns for neurological impairment (e.g. being SGA)]. Infants with major congenital anomalies or genetic or syndromic conditions were not included.

The study protocol received approval from the Ethics Committee for Medical Imaging Research of the CERF (“Collège des Enseignants en Radiologie de France”) and the “Comité de Protection des Personnes” Ile de France 3 (protocol DEVine, CEA 100 054). All procedures conformed to the principles of the Declaration of Helsinki and adhered to local data protection regulations.

### MRI Data Acquisition

MRI examinations were performed at TEA (post-menstrual age (PMA) between 37 and 43 weeks) following a uniform imaging protocol on a Philips Ingenia 3.0 Tesla scanner equipped with a 32-channel head coil and 45 mT/m gradient strength. No sedation was administered; infants were scanned during natural sleep following feeding, using immobilization and auditory protections to minimize motion and discomfort.

The imaging protocol included multiple sequences, in particular the following ones used for this study:

- T2-weighted (T2w) 2D images (turbo spin echo, TSE factor = 34) were acquired in coronal, axial, and sagittal orientations with parameters set as follows: echo time TE = 150 ms, repetition time TR = 5500 ms, slice thickness of 2.0 mm, flip angle of 90°, in-plane field of view FOV = 153 × 153 mm, reconstructed matrix size = 192 × 192, and in-plane voxel size = 0.8 × 0.8 mm². Parallel imaging was used with a SENSE factor of 2.8. Acquisition times were 2 minutes and 45 seconds or 3 minutes and 18 seconds for axial and sagittal planes (50 or 60 slices), and 3 minutes and 18 seconds or 3 minutes and 51 seconds for coronal plane (60 or 70 slices).
- DWI images were obtained with an echo-planar imaging (EPI) sequence, in the axial plane using a single-shell scheme with 42 diffusion gradient directions at b = 1000 s/mm², and four interleaved b=0 volumes. Imaging parameters were TE = 75 ms, TR = 5000 ms, FOV = 160 × 160 × 108 mm, reconstructed matrix 80 × 80, isotropic in-plane voxel size = 2 mm, 54 slices, thickness = 2mm, SENSE factor = 1.7; MB = 2; halfscan factor = 0.75. The phase-encoding direction was postero-anterior, and acquisition lasted approximately 4 minutes.
- An additional b=0 EPI sequence was acquired with similar parameters, for three volumes at b = 0 s/mm² and phase-encoding in the reverse (antero-posterior) direction for later susceptibility distortion correction. This sequence lasted approximately 25 seconds.

### MRI Processing

Processing of both T2w and DWI images was performed using a combination of state-of-the-art pipelines and tools optimized for neonatal brain imaging, which we specifically adapted to our infant population both with and without macroscopic brain lesions. The whole pipeline, with the successive steps detailed below, is summarized and illustrated in Figure 1.

**Figure 1.**
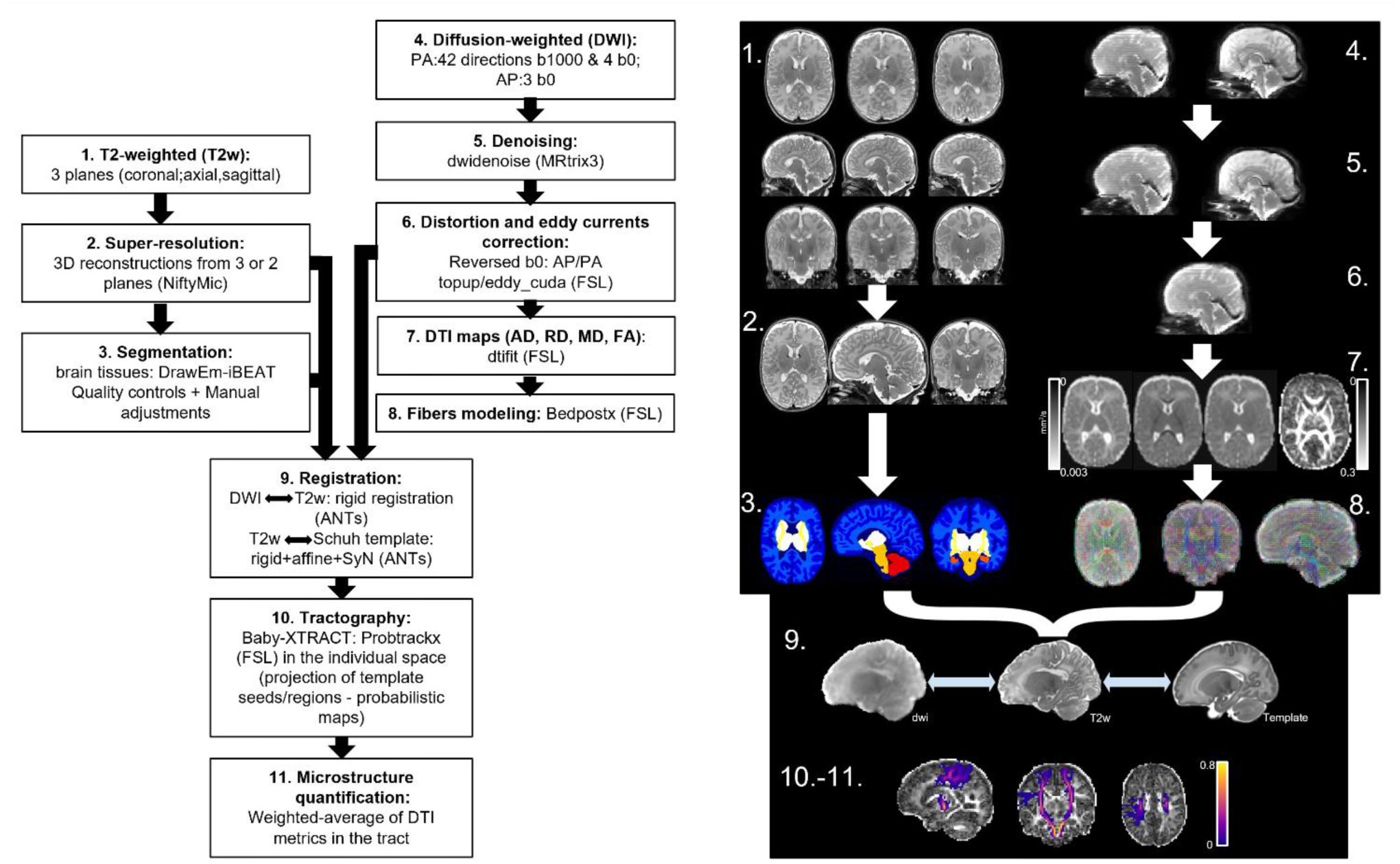
Diagram of the preprocessing pipeline. Left: Different steps taken and tools used. Right: An example of the different stages illustrated for a subject.

### T2w processing

The quality of raw MRI data was systematically assessed prior to preprocessing. Visual inspection was performed to identify major artifacts such as motion, ghosting, signal dropout or enhancement, and incomplete brain coverage. While most T2w images met quality standards, some were affected by excessive motion or other artefacts that compromised their usability. T2w anatomical images were first processed using a super-resolution algorithm to improve spatial resolution and enhance structural details. This was achieved using the NiftyMIC tool (Ebner et al., 2020), which reconstructs high-resolution 3D volumes from multiple complementary 2D stacks. In this study, the three T2-weighted acquisition planes (axial, coronal, and sagittal) were used when available and with sufficient quality. In cases where one plane was missing or severely affected by motion artifacts, reconstruction was performed using the remaining two planes. The resulting super-resolved images had an isotropic voxel size of 0.8 mm, and this was used in all the subsequent steps of processing.

Brain tissue segmentation and anatomical parcellation were conducted using a combination of tools to maximize accuracy across brain regions (Elbaz et al., 2025). The DrawEM pipeline from the *developing Human Connectome Project* (Makropoulos et al., 2018) was first used due to its strong performance on neonatal data and the availability of atlases for the considered PMA. However, for our preterm population, DrawEM tended to perform sub-optimally for cortical identification and at the interface with cerebellum. Thus, this approach was complemented with segmentation obtained with iBEAT pipeline (version 2.0) (Wang et al., 2023). This hybrid strategy (detailed in (Elbaz et al, 2025)) enabled high-quality parcellation of the whole brain, including more reliable labeling of infratentorial structures.

Besides, brain masks necessary for registration (details below) were generated using the Brain Extraction Tool (BET) from FSL (version 6.0.7.13) (Smith, 2002). When needed, parameters such as the fractional intensity threshold were manually adjusted to optimize brain extraction. All resulting masks were visually inspected to ensure accurate delineation of brain tissues before subsequent processing steps.

### DWI processing

DWI data underwent a rigorous preprocessing pipeline to ensure optimal data quality and minimize artifacts. Initial denoising was performed using the *dwidenoise* function from the MRtrix3 toolbox (version 3.0.3) (Tournier et al., 2019), which applies a method based on principal component analysis to reduce random noise.

Motion and susceptibility-induced distortions were corrected using the *dwifslpreproc* function (from MRtrix3; Tournier et al., 2019), which wraps FSL *topup* and *eddy_cuda* tools (Andersson et al., 2016). For this step, pairs of b0 images with reversed phase-encoding directions (across the 3 volumes for antero–posterior direction and the 4 volumes for the postero–anterior direction) were manually selected to optimize distortion correction.

A corrected b0 image, derived from the best quality manually selected b0 volume, was used to create the brain mask after preprocessing. The process followed the same approach as for T2w images (FSL BET), including manual parameter tuning when necessary and visual inspection to ensure accuracy before subsequent processing.

Moreover, for each subject, we estimated the DTI model with FSL (version 6.0.7.13, Behrens et al., 2003). DTI parameter maps were computed in native space for FA, MD, axial diffusivity (AD), and radial diffusivity (RD) (results for the latter two metrics are provided in Supplementary Information). In addition to their sensitivity to WM alterations, these metrics are greatly impacted by the progressive brain maturation and WM myelination, particularly during infancy: in most brain regions, FA increases with PMA at MRI, while diffusivities decrease (Ouyang et al., 2019). Therefore, we expected the variability in the cohort PMA (∼4 weeks) to potentially have an impact on the DTI metrics (see below for statistical analyses).

### Registration procedure

Spatial transformations were required to transfer anatomical priors from a template space to each subject’s individual DWI space, notably for the projection of regions required for the tractography procedure. For each transformation step, diffeomorphic registration methods were used, allowing bidirectional mapping from the template to individual T2w images and subsequently to DWI space, and vice versa. Because of the anatomical particularities of this preterm cohort due to brain lesions, the registration procedures were optimized specifically as detailed below, with adapted registration parameters and brain masks.

The T2w images were non-linearly registered to the neonatal template (Schuh et al., 2018) using a combination of rigid, affine, and nonlinear symmetric diffeomorphic transformations to standardize anatomical space across subjects (ANTs version 2.5.3) (Avants et al., 2011). To improve accuracy, brain masks derived from tissue segmentations were used to exclude cerebrospinal fluid (CSF) during the T2w-template alignment. This masking was crucial, as one of the most common sources of misregistration was the presence of sharp intensity transitions caused by hyperintense T2w signal in liquids, which frequently led to confusion between CSF and the cortical ribbon, or between CSF and the corpus callosum.

In parallel, DWI images were registered to the anatomical T2w images on a subject-specific basis using ANTs, employing a rigid registration to align diffusion space and T2w space. The composition of template-T2w and T2w-DWI transformations enabled direct warping from the template space to each subject’s individual DWI space.

### Reconstruction of WM tracts

#### Estimation of diffusion model

To model complex fiber architecture, fiber orientation distributions were estimated using FSL Bedpostx (Hernandez et al., 2013) with a single-shell ball-and-stick model and parameters set to two fibers per voxel, weight = 1 and a burn-in period of 1000 iterations, enabling robust Bayesian estimation of crossing fibers in neonatal data. Compared with the diffusion tensor model, this approach allowed a secondary fiber orientation to be retained when supported by the diffusion data (Behrens et al., 2007), and was more reliable for the estimation of tractography trajectories. The masks, derived from the optimal b0 volume and covering the entire brain, served as the spatial constraint for estimating the fiber orientation model.

#### Probabilistic tractography

To enable detailed and anatomically specific analysis of WM development in neonates, major WM tracts were identified using baby-XTRACT (Warrington et al., 2022), an automated and neonatal-specific tractography protocol, using FSL ProbtrackX. Baby-XTRACT includes segmentation protocols (Warrington et al., 2022) for 42 distinct WM tracts, subdivided into several categories: 5 projection tracts, 10 association fiber tracts, and 4 limbic system tracts, each represented bilaterally with separate left and right hemisphere reconstructions, as well as 4 commissural fiber tracts. It was performed in each subject’s native diffusion space, preserving individual anatomical features. The approach relies on anatomical regions (seeds, targets, waypoints, stops, and exclusions), defined in the Schuh template space. These regions were transformed into each subject’s native DWI space using the ANTs-derived registrations described above, with the composed template-to-T2w and T2w-to-DWI transformations. Fiber modelling and probabilistic tractography were then performed based on the native preprocessed DWI data, at a spatial resolution of 2mm isotropic.

#### Sensorimotor tracts

In the present study, we focused on tracts involved in sensorimotor functions, selecting a subset of 5 tracts as detailed in introduction: 3 bilateral tracts with the corticospinal tract (CST), superior thalamic radiation (STR) and frontal aslant tract (FAT); as well as two commissural tracts: the forceps minor (FMI) and middle cerebellar peduncle (MCP).

#### Individual-level quality control and tract optimization

The quality of the reconstructed tracts was visually verified in 25 selected subjects, representative of the clinical and brain anatomical variability (e.g., varying GA at birth; presence or absence of brain lesions of different types and severities; varying amounts of extra-axial CSF associated with more or less marked widening of the pericerebral subarachnoid spaces), and with various data quality (e.g., varying levels of motion during acquisition; different qualities of motion correction). This aimed to cover the main scenarios likely to influence the quality of tract reconstructions, and to ensure that the baby-XTRACT procedure, originally developed for full-term neonates (Warrington et al., 2022), remained reliable under varying conditions representative of the studied preterm cohort.

To reduce CSF-related partial-volume contamination of the tracts, subject-specific maps were masked for MD values lower than 0.0022 mm²/s: voxels with higher MD values were thus not considered in the extraction of tract-level DTI metrics, which aimed to improve the reliability of the quantification detailed below.

#### Group-level probabilistic tractography maps

To ensure the quality of the obtained tracts, we created probability maps showing the presence of each tract in each voxel at the group level (voxel-wise likelihood). This served two purposes: verifying the anatomical consistency of the tracts and observing the inter-individual variability for each tract. To compute these maps, individual tractography results were first thresholded at 10% of the maximum tract density to remove spurious connections, and then binarised to generate subject-specific presence masks. These binary masks were then transformed into the common neonatal template space (Schuh et al., 2018) and summed across subjects and converted to percentages, considering the number of subjects for which each tract was reconstructed (see Results section “Evaluation of the reconstructed WM tracts: Individual tractography”). These maps then represented the proportion of subjects in which each voxel was included in a given tract.

### Microstructure quantification

Once the tracts were obtained in the individual native space, DTI metrics (FA, MD, AD, RD) were extracted for each tract and each subject, by computing a weighted average (Adibpour et al., 2018). This accounted for the probabilistic nature of tractography, by weighting each tract voxel by the number of streamlines in this voxel, using the relative probability map of the tract provided by ProbtrackX (i.e., densityNorm map representing streamline density normalized by the number of seeds). This weighting provided greater importance to the core of the tract, which is more reliably reconstructed across subjects, than to its peripheral regions that are less consistent and more variable.

For bilateral tracts involved in sensorimotor functions, DTI metrics for the left and right hemispheres were further averaged, because studying potential inter-hemispheric asymmetries was beyond the scope of this study and we had no specific hypotheses on the relationships between asymmetries and clinical factors. When an individual tract was not reconstructed in one hemisphere, the subject was not considered in the subsequent analysis for this tract.

Moreover, to obtain a global measure of the whole WM microstructure and integrity, we computed the averaged value of each DTI metric across the 42 tracts provided by the baby-XTRACT pipeline (Warrington et al., 2022). All reconstructed tracts were considered, even if a bilateral tract was successfully extracted only in one hemisphere. In this way, a “global WM metric” could be obtained for each subject.

### Clinical risk factors

Clinical information, including perinatal characteristics and the presence of major neonatal complications, was retrospectively extracted from the hospital’s electronic medical record system for analysis. As priorly detailed, we focused on 9 relevant clinical characteristics (Elbaz et al., 2025): sex, monochorionic twin pregnancy, category of GA at birth, being SGA at birth, the occurrence of BPD or NEC, the use of invasive mechanical ventilation or prolonged parenteral nutrition, and perinatal sepsis.

GA at birth was determined from first-trimester ultrasonography, and infants were grouped into three categories: <26 weeks (G1), 26-27weeks+6days (G2), and >=28weeks (G3). We initially planned to restrict G3 to infants born before 32w GA, but due to the uncertainty in estimating the exact date of conception, we decided to include two subjects who were supposed to be born at 32.4 weeks and 32.8 weeks GA and who presented typical lesions of prematurity (IVH 3). SGA classification was based on a birth weight below the 10th percentile for GA using sex-specific reference charts (AUDIPOG, 2025).

BPD was defined as the need for oxygen therapy beyond 36 weeks PMA or for more than 28 days postnatally. Invasive mechanical ventilation was considered if it exceeded a duration of 24 hours. NEC was diagnosed using a combination of clinical and radiological criteria in accordance with Bell’s classification (Bell et al., 1978), and we only considered stage II or higher. Prolonged parenteral nutrition referred to intravenous nutrition maintained for over 21 days. Sepsis was defined as any clinical suspicion of infection requiring an antibiotic treatment lasting more than five days, regardless of culture results.

To characterize the distribution of clinical risk factors across the groups of GA at birth, comparisons were performed using chi-square or Fisher’s exact tests, as appropriate. The resulting p-values were adjusted for multiple comparisons using the false discovery rate (FDR) procedure.

### Radiological severity scores

The Kidokoro scores were quantified for each infant based on structural MRI at TEA, as described in a previous study (Elbaz et al., 2025). This radiological scoring system is a standardized tool developed to assess brain abnormalities in preterm infants (Kidokoro et al., 2013). In the present study, we focused exclusively on the WM subscore, which quantifies specific abnormalities such as cystic lesions, focal signal abnormalities, delayed myelination, ventricular dilatation, corpus callosum thinning and reduced biparietal diameter (Elbaz et al., 2025). The Kidokoro WM scores range from 0 to 17 and were categorized into four levels of abnormalities: normal (0–2), mild (3–4), moderate (5–6), and severe (≥7).

### Statistical analysis

We aimed to assess whether the inter-individual variability in DTI metrics in the global WM and in each tract could be explained by a set of clinical factors. Results for the different tracts were compared in a descriptive and qualitative way. Statistical tests were chosen based on data distribution and model assumptions, and multivariate linear models including both categorical and continuous factors appeared to be the most adequate to answer our questions. The main assumptions of the statistical models were scrupulously verified beforehand, including linearity, approximate normality of residuals, homoscedasticity, and the absence of problematic multicollinearity among predictors. Multicollinearity was evaluated using variance inflation factors (VIF). When initial model diagnostics were not satisfactory for certain tract– metric combinations, influential observations were examined using Cook’s distance (outlier threshold = 4/n, n defined as the number of subjects for this specific tract). Cook’s distances were computed separately for each tract, and only for the tracts and metrics that violated the model assumptions; but when a subject was identified as an outlier for a certain tract-metric combination, the subject was removed for all metrics in that tract. The corresponding models were then refitted after exclusion of influential observations, and all diagnostic criteria were reassessed. This led to a varying number of considered subjects across analyses. Model fit was summarized using R² and adjusted R².

First, we modelled each DTI metric for the global WM and each tract separately as a function of PMA at MRI (continuous variable) and categorized risk factors (9 clinical factors and the class of WM injury severity derived from the Kidokoro score). Second, each tract-specific DTI metric was modelled as a function of the global WM metric and the same risk factors. This allowed us to include global WM integrity and maturation as a regressor of no interest, thereby disentangling effects reflecting overall WM involvement from those more specifically attributable to the tract of interest. Third, a complementary set of analyses was performed after restricting the sample to infants with normal Kidokoro scores and without macroscopically detectable lesions (i.e., without IVH grades II-III-IV). This aimed to homogenize the population to better estimate the effects of clinical factors on the tract microstructure within little-injured WM.

The reference category for each categorical factor was defined *a priori* as the lowest-risk condition (e.g., female for sex; not being SGA). For GA at birth, group G2 (and not G3) was selected as the reference category to better take account of the GA effect: indeed, infants in G3 were scanned because a neurological abnormality was suspected (which was not the case for G2).

For all analyses, statistical significance was set at p < 0.05 after correction for multiple comparisons across tracts. We applied this correction with FDR method, separately for each DTI metric. Corrected p-values below 0.10 are also reported as statistical trends. All analyses were performed using R software version 4.4.0 (R Core Team, 2025).

## Results

### Cohort description and quality control of MRI data

A total of 111 preterm infants were included, and their clinical characteristics are detailed in Table 1 (GA at birth: mean = 27.6 weeks, standard deviation SD = 2.05, [24.1w; 32.9w]; PMA at MRI examination: mean = 41.3 weeks, SD = 0.65, [38.4w; 42.6w]. A wide variety of brain characteristics and abnormalities was observed across the cohort (Figure 2).

**Figure 2.**
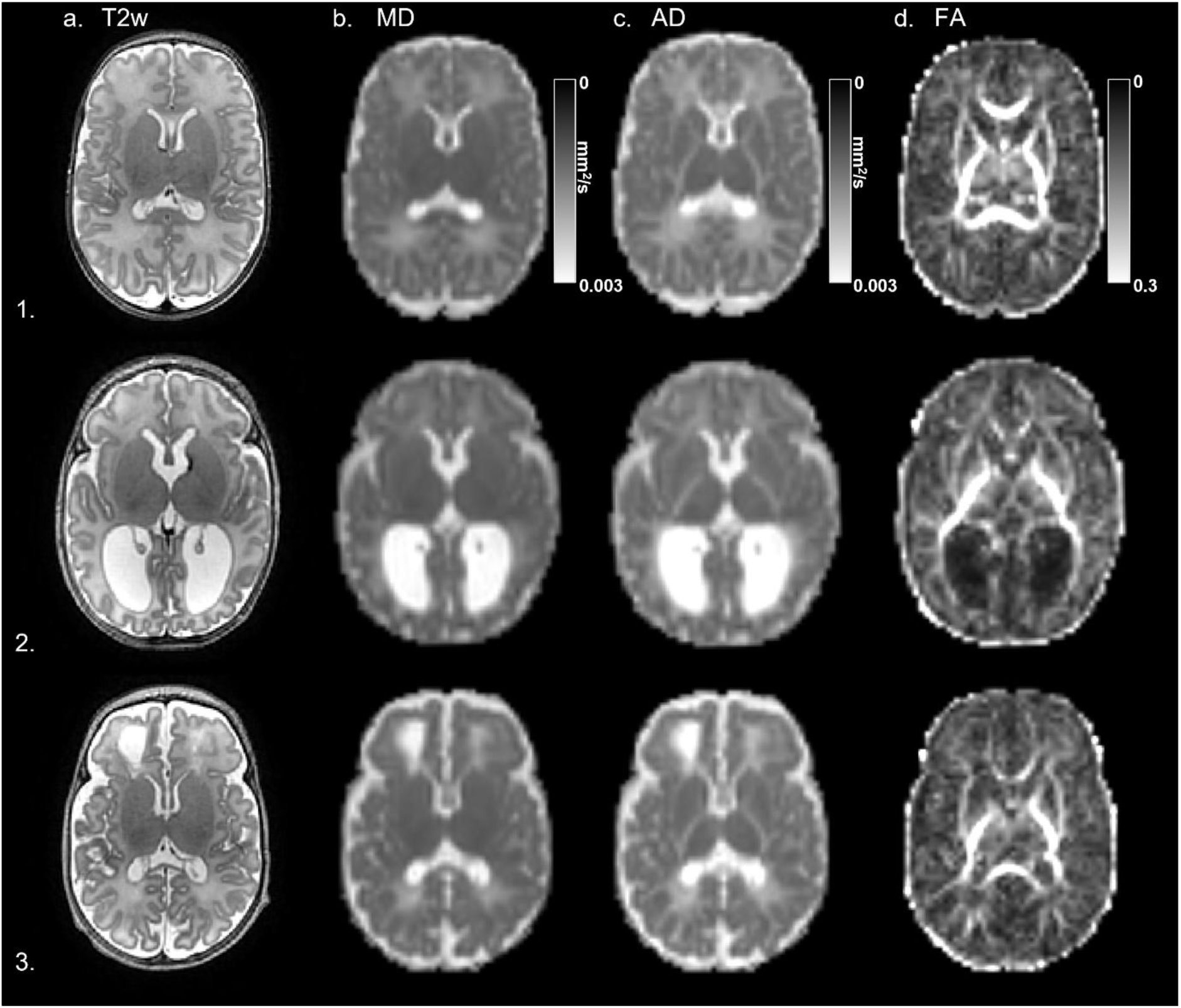
Anatomical MRI and DTI images of three representative infants. Images are presented on an axial slice at the level of the lateral ventricles. From left to right: (a) T2-weighted anatomical images showing structural brain anatomy. (b) Mean diffusivity (MD) map indicating the average diffusivity of water molecules, with increased values in CSF and reduced diffusivity in tissues with high cellularity (e.g. cortex). (c) Axial diffusivity (AD) map reflecting the diffusion of water parallel to axonal fibers, typically high in tightly packed white matter regions (e.g. internal capsules). (d) Fractional anisotropy (FA) map showing anisotropic diffusion, with high values in regions of fibers organized in dense tracts (e.g. corpus callosum). Radial diffusivity (RD) maps were omitted for clarity due to their high redundancy with other diffusivity metrics. From top to bottom: (1) Infant without evidence of cerebral lesions or abnormalities. (2) Infant with ventricular dilatation following grade 3 intraventricular hemorrhage (IVH3) and bilateral subependymal hemorrhages. (3) Infant presenting diffuse WM signal abnormalities in frontal lobe with cyst in the context of cavitary periventricular leukomalacia (PVL).

**Table 1.**
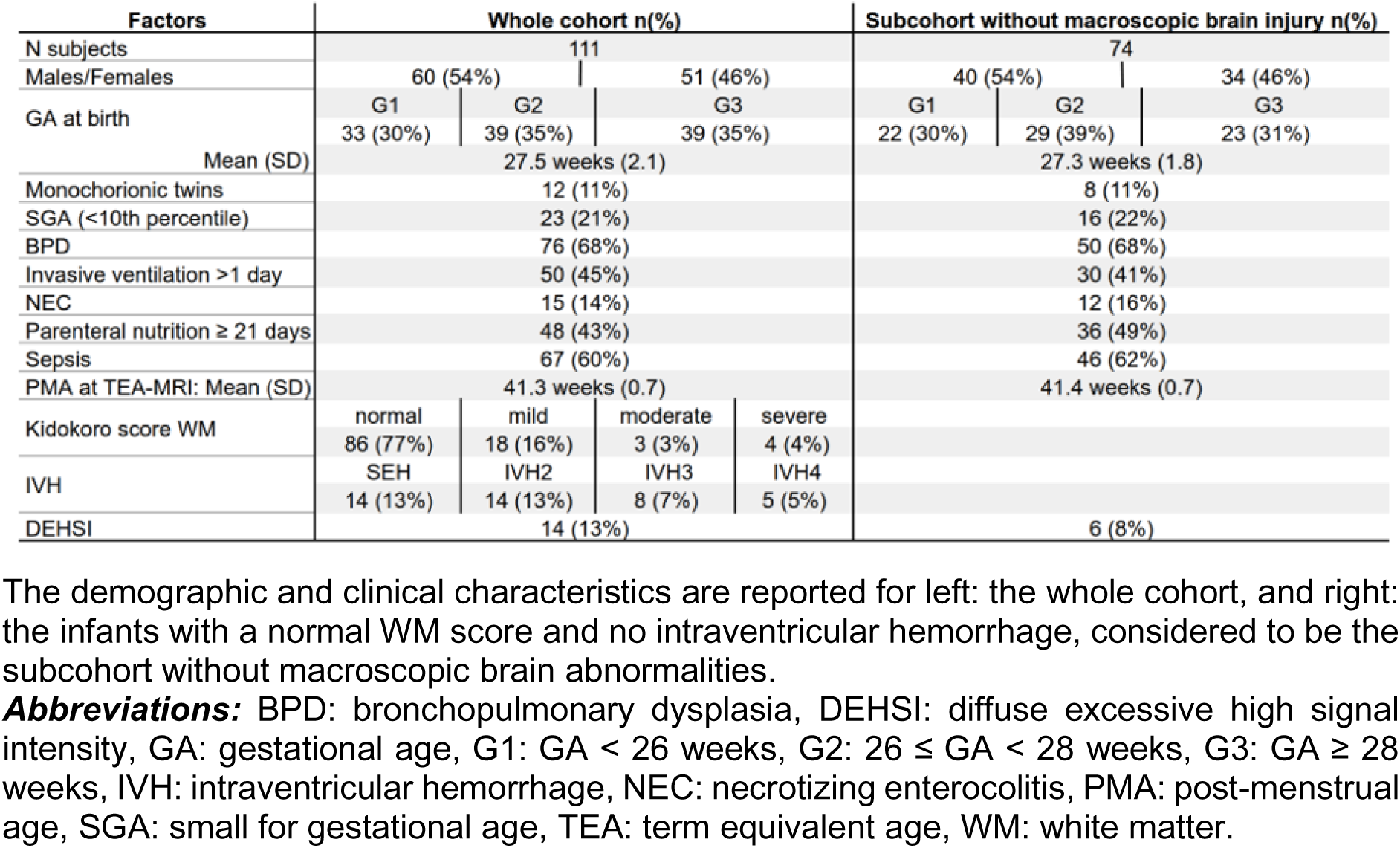
Characteristics of the cohort of premature-born infants.

| Factors | Whole cohort n(%) |  |  |  | Subcohort without macroscopic brain injury n(%) |  |  |
| --- | --- | --- | --- | --- | --- | --- | --- |
| N subjects | 111 |  |  |  | 74 |  |  |
| Males/Females | 60 (54%) |  | 51 (46%) |  | 40 (54%) |  | 34 (46%) |
| GA at birth | G1 | G2 | G3 |  | G1 | G2 | G3 |
|  | 33 (30%) | 39 (35%) | 39 (35%) |  | 22 (30%) | 29 (39%) | 23 (31%) |
| Mean (SD) | 27.5 weeks (2.1) |  |  |  | 27.3 weeks (1.8) |  |  |
| Monochorionic twins | 12 (11%) |  |  |  | 8 (11%) |  |  |
| SGA (<10th percentile) | 23 (21%) |  |  |  | 16 (22%) |  |  |
| BPD | 76 (68%) |  |  |  | 50 (68%) |  |  |
| Invasive ventilation >1 day | 50 (45%) |  |  |  | 30 (41%) |  |  |
| NEC | 15 (14%) |  |  |  | 12 (16%) |  |  |
| Parenteral nutrition ≥ 21 days | 48 (43%) |  |  |  | 36 (49%) |  |  |
| Sepsis | 67 (60%) |  |  |  | 46 (62%) |  |  |
| PMA at TEA-MRI: Mean (SD) | 41.3 weeks (0.7) |  |  |  | 41.4 weeks (0.7) |  |  |
| Kidokoro score WM | normal | mild | moderate | severe |  |  |  |
|  | 86 (77%) | 18 (16%) | 3 (3%) | 4 (4%) |  |  |  |
| IVH | SEH | IVH2 | IVH3 | IVH4 |  |  |  |
|  | 14 (13%) | 14 (13%) | 8 (7%) | 5 (5%) |  |  |  |
| DEHSI | 14 (13%) |  |  |  | 6 (8%) |  |  |

The demographic and clinical characteristics are reported for left: the whole cohort, and right: the infants with a normal WM score and no intraventricular hemorrhage, considered to be the subcohort without macroscopic brain abnormalities.
**Abbreviations:** BPD: bronchopulmonary dysplasia, DEHSI: diffuse excessive high signal intensity, GA: gestational age, G1: GA < 26 weeks, G2: 26 ≤ GA < 28 weeks, G3: GA ≥ 28 weeks, IVH: intraventricular hemorrhage, NEC: necrotizing enterocolitis, PMA: post-menstrual age, SGA: small for gestational age, TEA: term equivalent age, WM: white matter.

### Clinical characteristics

Some co-occurrences of neonatal morbidities and treatment-related exposures were observed across infants (see Sup.Figure 1 for pairwise association matrix between clinical variables), such as associations between respiratory complications (invasive mechanical ventilation, BPD), prolonged nutritional nutrition, necrotizing enterocolitis and sepsis. An association between GA group and SGA status was also observed. While not included in the statistical models, we noted that intracranial volume was associated with SGA status and invasive mechanical ventilation. These associations justified the need for careful evaluations of multicollinearity in the statistical models described below.

The distribution of clinical characteristics across groups of GA at birth is further detailed in Sup.Table 1. After FDR correction, significant between-group differences were observed for monochorionic twin pregnancy, SGA status, BPD, invasive mechanical ventilation, prolonged parenteral nutrition, and sepsis. Actually, G1 infants most frequently required invasive mechanical ventilation and had the highest frequencies of prolonged parenteral nutrition and sepsis. Conversely, G3 infants were less frequently affected by BPD, prolonged parenteral nutrition, and sepsis, but showed the highest proportion of monochorionic twin pregnancy and SGA status. Besides, the frequency of NEC did not differ significantly between groups.

### Kidokoro WM subscore

The radiological WM characteristics were classified as normal for 86 infants (77.5%), whereas the severity of abnormalities was classified as mild for 18 (16.2%), moderate for 3 (2.7%), and severe for 4 (3.6%). 74 infants (66.7%) were considered to have no macroscopic abnormalities (normal WM Kidokoro score and no significant brain lesion).

### T2w data

While most T2w images met quality standards, the quality of one of the three planes was insufficient for 33 out of 111 subjects (30%). Then only two motion-free planes were used to reconstruct the 3D anatomical volume with the super-resolution approach. All the resulting super-resolved T2w images showed sufficient quality for further processing and analyses. Tissue segmentation maps generated using iBEAT and DrawEM were systematically reviewed through visual quality control. Manual corrections were performed in about 25% of segmentations, in cases of visible segmentation errors mainly consisting of (i) mislabeling of tissue within the CSF, particularly in the presence of large pericerebral spaces, and (ii) difficulties in the identification of the cerebellum with both methods (Elbaz et al., 2025).

### DWI data

Among the 111 DWI datasets acquired, the majority were of high quality, with fewer than 10 motion-corrupted volumes (out of 42 volumes at b=1000) and without brain cropping. Scans of 6 infants (5.4%) exhibited more significant quality issues, either due to motion artifacts or partial brain exclusion during acquisition. Nevertheless, after preprocessing steps (including denoising, motion correction, eddy current correction, and distortion correction), all DWI datasets were considered of sufficient quality for further analyses, as the remaining issues were limited to minor cropping in the very inferior part of the cerebellum (tonsils and inferior surface) or a very small portion of the fronto-parietal cortex, none of which involved the tracts analysed in this study

### Evaluation of the reconstructed WM tracts

#### Individual tractography

Considering all the 42 tracts generated by baby-XTRACT, the variability in reconstruction was quite important (Sup.Table 2). 5 tracts were particularly difficult to reconstruct because of their small size in the infant brain or their well-known inter-hemispheric asymmetries (Dubois et al., 2016): the arcuate fasciculus and acoustic radiations mostly in the right hemisphere, the superior longitudinal fasciculus II mostly in the left hemisphere, the fornix and the cingulum peri-genual subsection on both sides.

**Table 2.**
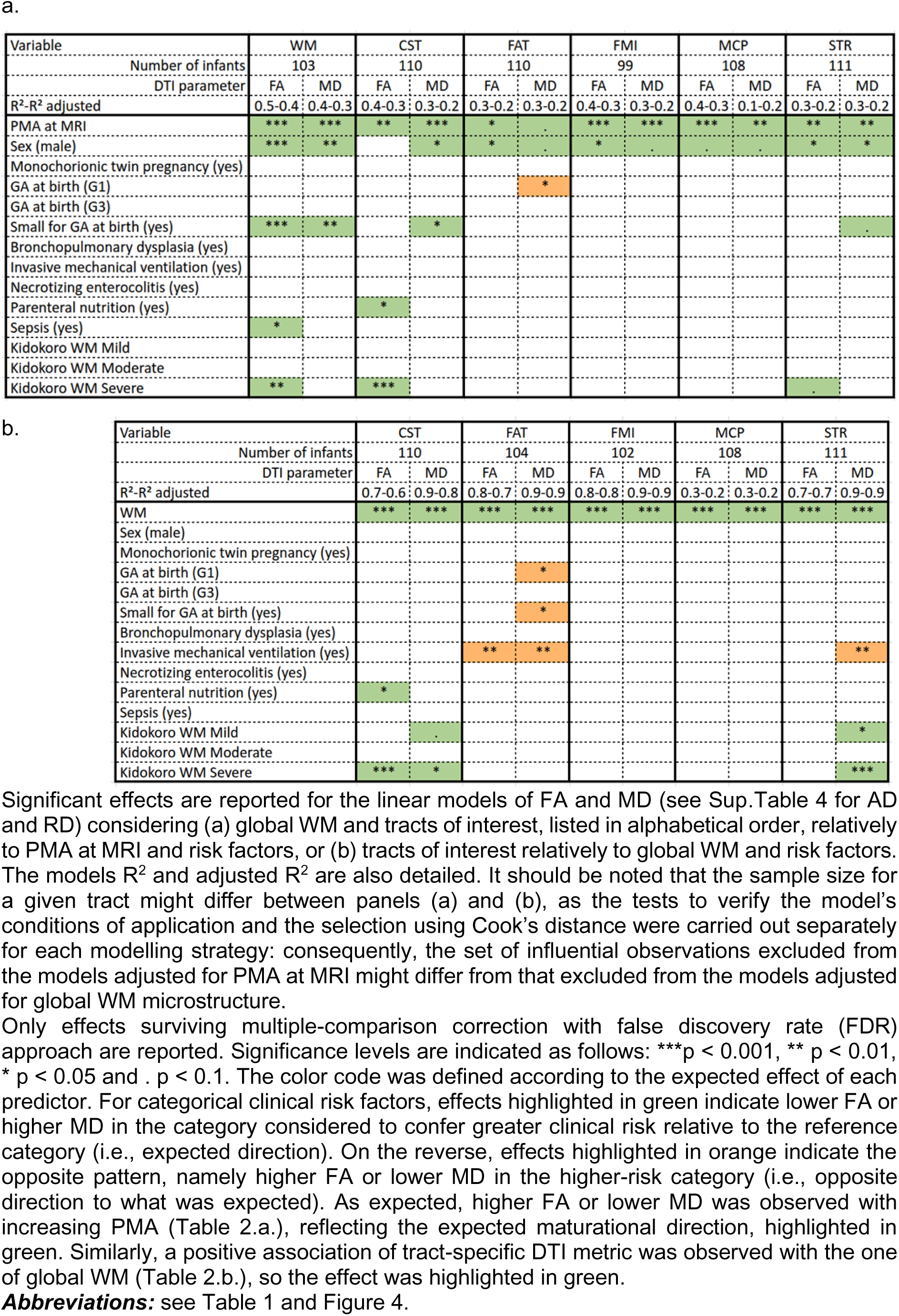
Summary of the results of statistical analyses over the whole cohort for fractional anisotropy (FA) and mean diffusivity (MD)

All 3 bilateral sensorimotor tracts were successfully reconstructed in both hemispheres in the vast majority of the 111 subjects: 110 infants for CST, 111 for STR, 110 for FAT. The commissural tracts were also successfully reconstructed over the cohort, for 111 infants for FMI and 108 for MCP. As detailed in Sup.Table 3, causes for these unsuccessful tract reconstructions included lesions along the expected course of the tract, a registration issue, or no clear anatomical or technical reason. Visually checking the trajectory of individual tracts for 25 subjects revealed the high quality of reconstructions, although some inter-individual variability was observed across infants.

**Table 3.**
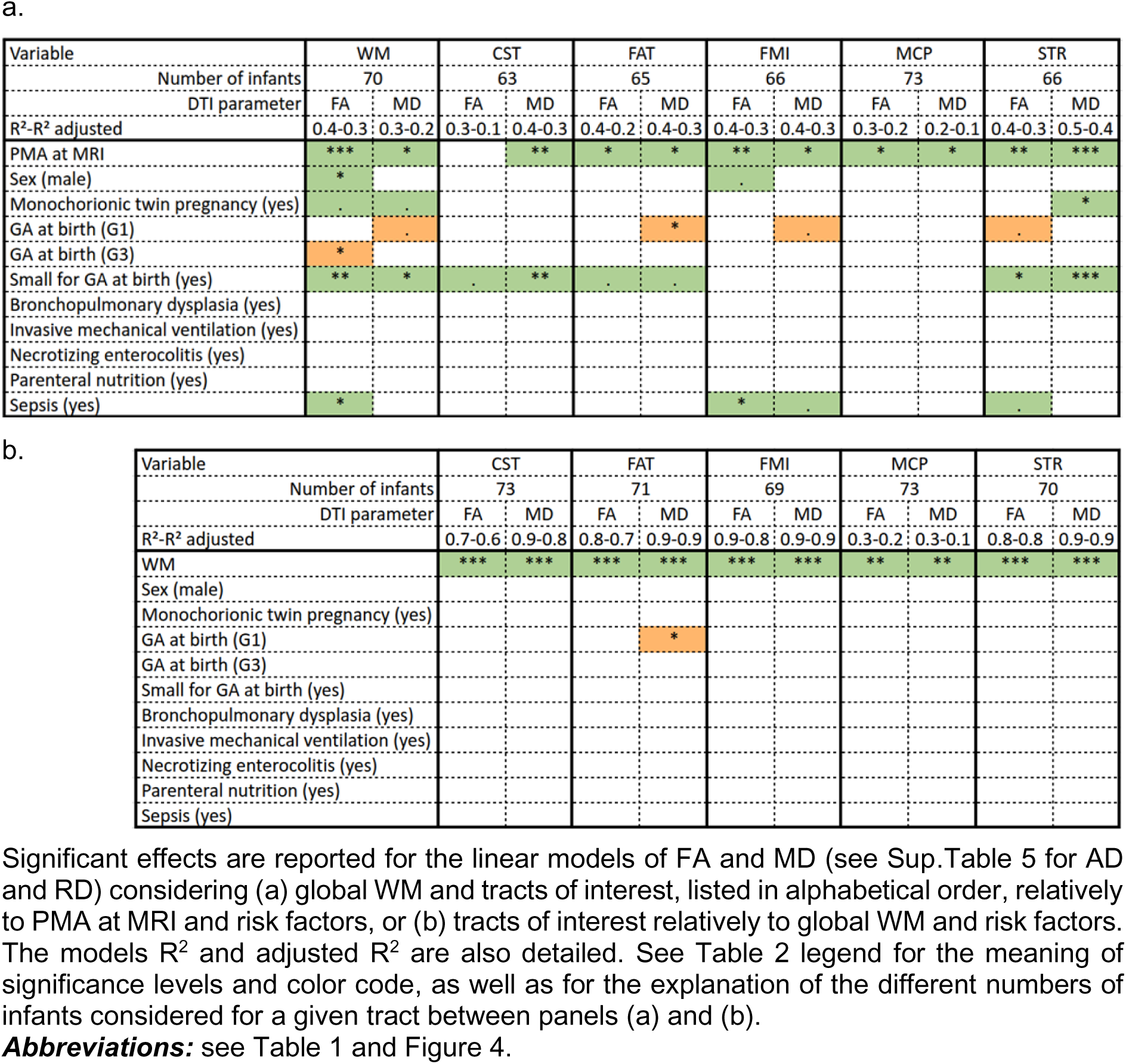
Summary of the results of statistical analyses over the sub-group of infants without macroscopic brain abnormalities for FA and MD.

### Tractography maps over the infant group

Group-level probabilistic maps then helped us to verify the tract quality over the group and assess the consistency and anatomical plausibility of trajectories. Results were in line with known neuroanatomy in adults and in newborns without cerebral injury (Figure 3).

**Figure 3.**
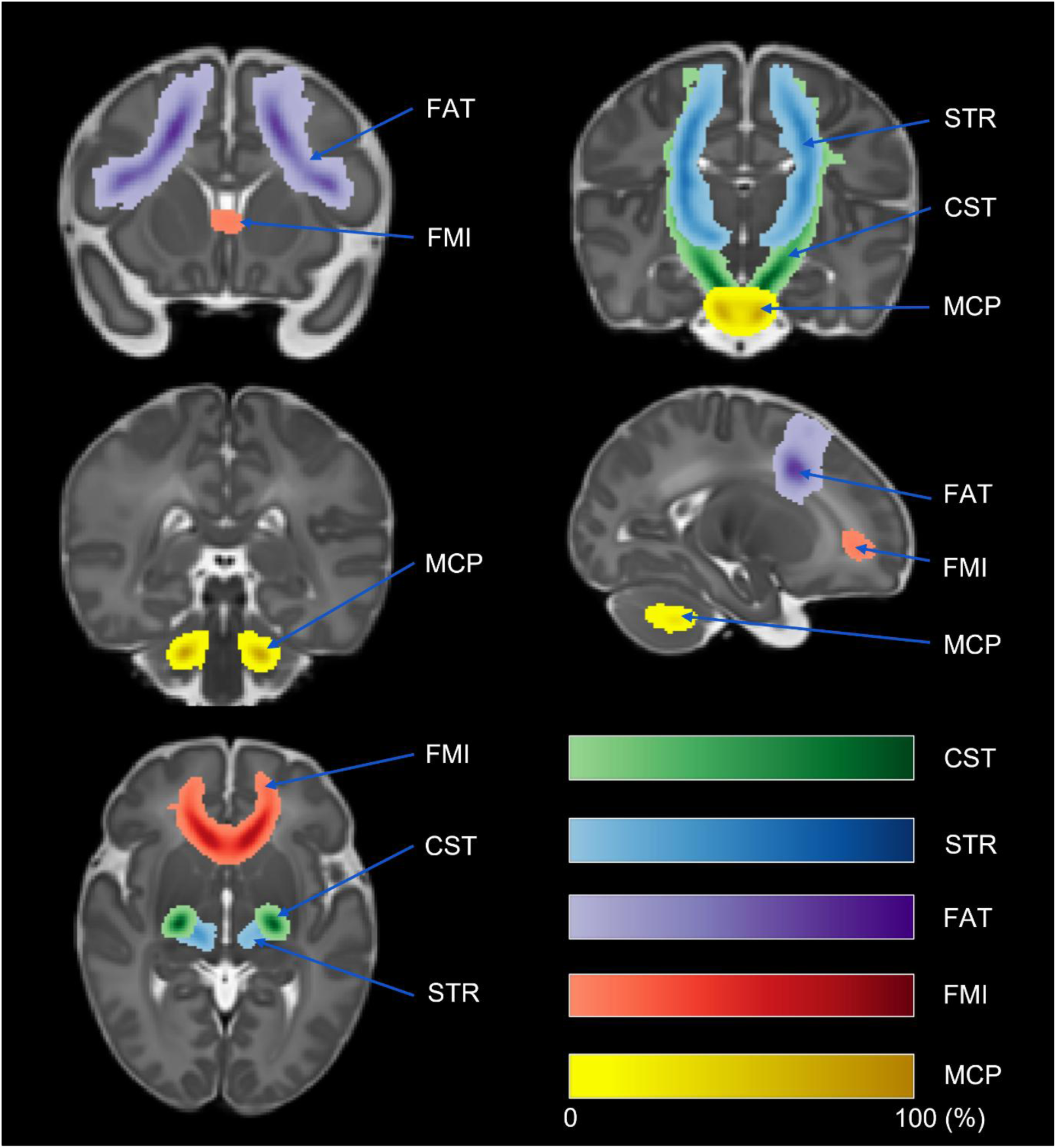
Group-level probabilistic maps of the tracts of interest. These maps show the voxelwise probability of tract presence across the entire cohort, quantified as the percentage likelihood (0–100%) of each white matter pathway. Coronal, sagittal, and axial planes, illustrating the 5 tracts of interest, involved in sensorimotor network (with the number of infants with reconstructed tracts): 3 bilateral bundles: the corticospinal tracts (CST, N=110), superior thalamic radiations (STR, N=111) and frontal-aslant tracts (FAT, N=110); as well as 2 commissural tracts: forceps minor (FMI, N=111) and middle cerebellar peduncle (MCP, N=108), all projected on the Schuh template.

### Exploration of the inter-individual variability in the microstructure of global WM and tracts

#### Quantification of WM microstructure

First, although some of the 42 tracts failed to be reconstructed for some infants (Sup.Table 2), this did not prevent the computation of DTI metrics for the global WM, and the resulting quantification was not particularly skewed as these subjects were not detected as outliers in the following analyses on global WM. At the group level, DTI metrics revealed quantitative differences across the 5 tracts of interest, reflecting the expected diversity of microstructural characteristics at TEA, e.g., FA was the highest in the CST and the lowest in FAT, while MD was lower in CST, STR and MCP than in FAT and FMI (Figure 4 for FA and MD. See Sup.Figure 2 for AD and RD). A wide inter-individual variability in DTI metrics was further observed. To investigate the potential impact of clinical risk factors on the microstructure of global WM and the 5 tracts of interest, different statistical models were conducted.

**Figure 4.**
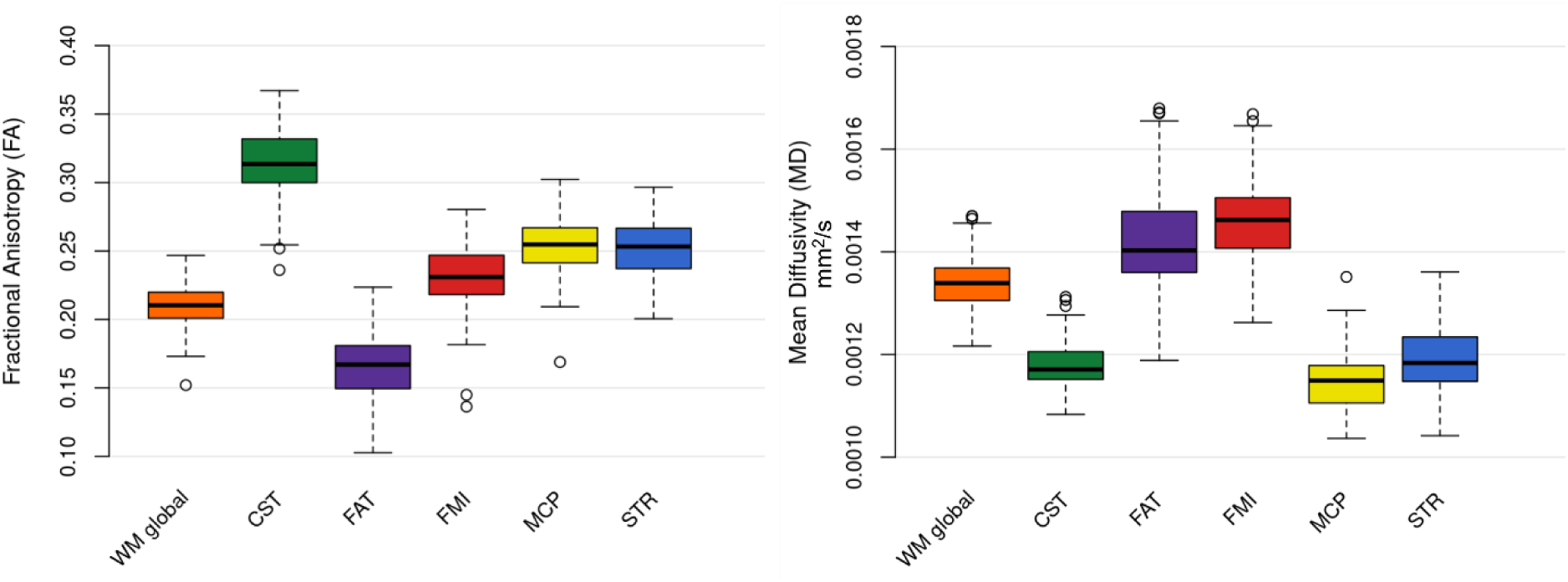
Quantification of the white matter microstructure with fractional anisotropy (FA) and mean diffusivity (MD) Box plots of the weighted mean values of FA (left) and MD (right) parameters quantified in the global white matter (42 tracts) and in the 5 tracts of interest, listed in alphabetical order. ***Abbreviations:*** CST: corticospinal tract, FAT: frontal-aslant tract, FMI: forceps minor, MCP: middle cerebellar peduncle, STR: superior thalamic radiation, WM: white matter.

### Models accounting for PMA at MRI

The linear models on FA and MD of global WM first highlighted significant effects of PMA at MRI, sex, being SGA at birth, sepsis and Kidokoro severe class (Table 2a, Sup.Figure 3a. See Sup.Table 4 for results for AD and RD). More specifically, as expected, FA increased with PMA at scan while MD decreased. Being a male or SGA was associated with lower FA values and higher MD values. Having sepsis or a severe Kidokoro score was associated with lower FA values.

When this analysis was restricted to the sub-cohort of infants without macroscopic brain abnormalities (Table 3a), similar findings were observed for PMA at MRI, sex, being SGA at birth and sepsis. We further observed a trend for alterations associated with monochorionic twin pregnancy (lower FA, higher MD), as well as some impact of GA at birth (lower MD in G1 than G2) but in the opposite direction to our hypotheses, which was therefore unexpected.

The linear models run for the 5 tracts of interest over the whole cohort also revealed main effects of PMA at MRI and sex on FA and MD (Table 2a, Sup.Figures 3b, 3c). For the CST, we further observed that being SGA at birth was associated with higher MD, and parenteral nutrition and WM abnormality severity were associated with lower FA. Trends for effects of SGA status and Kidokoro severity were also observed for STR. Surprisingly, in FAT, lower MD was observed in G1 than in G2 group.

In the sub-cohort of infants without macroscopic brain abnormalities (Table 3a), the models confirmed consistent effects of PMA at MRI on FA and MD in all tracts. The effect of sex was only observed as a trend in FMI. Being SGA had an effect on CST, STR and FAT characteristics. The STR microstructure was related to monochorionic twin pregnancy and to sepsis that also impacted the FMI. An unexpected impact of GA at birth was observed for STR, FAT and FMI.

### Models accounting for global WM microstructure

In a second step, we aimed to investigate whether the impact of risk factors on WM tracts survived even when accounting for inter-individual differences in global WM integrity. We thus implemented models of DTI metrics including global WM (instead of PMA at MRI) as a covariate of non-interest that appeared to be associated with all tracts, as expected (Tables 2b, 3b, Sup.Figure 4). Different patterns of microstructure relationships with risk factors were then observed across tracts when considering the whole cohort (Table 2b). The microstructure of CST, and to a lesser extent of STR, were associated with prolonged parenteral nutrition needs and severity of the radiological WM abnormalities in expected ways. On the reverse, FAT showed an unexpected impact (lower MD +/- higher FA in at-risk conditions) of GA at birth (in G1), SGA and mechanical ventilation. This latter effect was also observed for STR. No significant associations were observed for FMI and MCP. When the sub-cohort of infants without macroscopic brain abnormalities was considered (Table 3b), only MD in FAT still showed the unexpected impact of GA at birth (in G1).

## Discussion

This study presents a detailed investigation of the impact of perinatal factors on WM microstructure within the sensorimotor network in extremely-to-very preterm infants scanned at TEA, using a dedicated processing pipeline of diffusion MRI and tractography. We observed both widespread dependence to some clinical risk factors (i.e., male sex, SGA, lesion severity and some neonatal morbidities) and heterogeneous patterns across tracts. In particular, the CST and FAT showed quite different profiles, in line with the hypothesis that different pathways may exhibit diverse vulnerability to prenatal contexts and perinatal exposures due to their specific developmental trajectories.

### Impact of prematurity-related risk factors on the WM microstructural maturation

Previous diffusion MRI studies have shown that multiple prematurity-related factors, including neonatal morbidities, impaired fetal or postnatal growth, and low birth weight, are associated with a global dysmaturation of WM microstructure, characterized by widespread abnormalities (Kaur et al., 2014; Lepomäki et al., 2013; Batalle et al., 2017). One of our objectives was therefore to distinguish the impact of clinical factors on the whole WM from the effects across the different tracts.

To begin with, it should be noted that neonatal morbidities and treatment-related exposures often co-occur in preterm cohorts, particularly among the most clinically vulnerable infants. In this study, we observed such co-occurrence to some extent (Sup.Figure 1). Associations between clinical factors might reflect shared underlying clinical vulnerability. For instance, invasive ventilation and BPD are variables characterizing a more severe and prolonged respiratory course. Their associations with prolonged parenteral nutrition likely reflect that infants requiring sustained respiratory and intensive care support also tend to achieve full enteral feeding later. Sepsis associations with prolonged parenteral nutrition and BPD may highlight the convergence of conditions among infants with greater immaturity and a more severe or prolonged neonatal course. Association between NEC and invasive mechanical ventilation may similarly reflect overlapping clinical severity. Therefore, neonatal risk factors co-occurred in this preterm cohort, and could not be regarded as biologically independent.

Infants of the three GA groups also differed in clinical characteristics (Sup.Table 1), with higher clinical vulnerability in G1 (higher proportion of infants with invasive mechanical ventilation, prolonged parenteral nutrition and sepsis) and less vulnerability in G3 (lower proportion of infants with BPD, prolonged parenteral nutrition, and sepsis). Furthermore, G3 showed the highest proportion of monochorionic twin pregnancy and SGA status, which might have led to the association between GA group and SGA status observed over the whole cohort. This was consistent with the indication-based inclusion of infants in this group, for whom MRI was performed because of the presence of clinical risk factors or suspected brain abnormalities. Despite these GA group differences, analyses stratified by group with greater homogeneity were not performed because of insufficient sample size and thus power.

Although some associations were observed between clinical factors, variance inflation factors (VIF) did not indicate problematic multicollinearity when pre-specified clinical factors were entered simultaneously into the statistical multivariate models of WM microstructure. This then appeared to be the most appropriate way to analyze the data. Consequently, associations attributed to individual factors should be interpreted as conditional markers of vulnerability rather than as isolated causal effects. As detailed below, adjustments for global white-matter microstructure further assessed whether these associations were relatively tract specific, but this was not intended to resolve the clinical dependence among predictors.

First, we observed that the **global WM** of preterm infants at TEA is affected by different clinical risk factors. A major effect of PMA at MRI was observed, in line with previous studies (e.g. Kimpton et al., 2021; Ling et al., 2013). Notably, an interval as short as 4 weeks seemed to modulate the WM microstructure, consistently with the observation that this period of brain development is particularly critical for this tissue growth and myelination (Kostović et al., 2019). A systematic dependence on sex was also demonstrated for global WM and all tracts, with boys showing a globally less mature WM microstructure than girls. This is also consistent with the well-established male vulnerability in preterm cohorts (Pierrat et al., 2021; Pogribna et al., 2013). Our findings further indicated that being SGA at birth is a significant risk factor for WM dysmaturation. This is in line with previous studies showing that SGA at birth and fetal growth restriction impact brain development at several levels and over long term, being associated with global and persistent alteration of WM maturation including widespread reductions in structure volumes and cortical surface area during childhood and adolescence (De Bie et al., 2011), and/or with WM microstructural alterations including reduced integrity and disrupted tract organization (Eikenes et al., 2012).

Interestingly, these effects of PMA, sex, and SGA on whole WM remained significant in the subgroup of preterm infants without macroscopic brain abnormalities. Based on additional targeted linear regression models including PMA at MRI, sex, SGA, macroscopic abnormality status (yes/no) and the corresponding two-way interactions (PMA-, sex- and SGA-by-abnormality status), we further observed that the associations of whole WM microstructure with PMA, sex, and SGA did not differ between infants with and without macroscopic brain abnormalities (results not shown). This supports the hypothesis that WM alterations are not limited to visible lesions but may reflect diffuse maturational disturbances affecting the whole WM. Widespread WM microstructural abnormalities have been previously reported at TEA in preterm infants without focal brain injury, including reduced FA across multiple WM regions (Anjari et al., 2007; Rose et al., 2013).

Also, neonatal sepsis effect on global WM microstructure is in line with previous literature associating sepsis with systemic inflammatory processes and/or WM microstructural alterations in very preterm infants at TEA (Anderson et al., 2024; Shim et al., 2012), as well as with poorer long-term motor outcomes and broader neurodevelopmental impairment (Hentges et al., 2013, Mitha et al., 2013; Cai et al., 2019). By contrast, beyond its association with global WM microstructure, the associations we found between WM microstructure and sepsis were limited to a small number of tracts. This might suggest that clinical sepsis may be by itself an insensitive indicator of the intensity and duration of the systemic inflammation underlying WM injury. Quantitative inflammatory biomarkers, such as C-reactive protein (CRP) or cytokine panels, would capture the inflammatory burden driving tract-level WM microstructural alterations more accurately, but these could not be collected due to limited blood sampling.

Beyond global WM alterations, we aimed to investigate **tract-specific vulnerability** by evaluating the differential sensitivity to risk factors across the tracts of the sensorimotor network. However, when tracts were analyzed using the same modelling strategy as global WM (i.e. adjusting only for PMA), few tract-specific effects emerged. An effect of GA group on FAT microstructure was observed, with infants born <26 weeks (G1) showing lower MD than those born at 26–28 weeks (G2). This pattern of association was unexpected, as G2 infants would typically be considered at relatively lower neurological risk than G1 infants and were thus expected to show more mature WM microstructure. Regarding parenteral nutrition, it was also associated with CST microstructure within our entire cohort. Previous studies in preterm infants have reported associations between early nutritional exposure, including parenteral nutrient intake, and WM maturation using ROI-based or TBSS diffusion MRI approaches (Schneider et al., 2018; Tam et al., 2016), but not with tract-specific investigations relying on tractography. This particular result is biologically plausible given the early maturation and vulnerability of the CST and the established influence of early nutritional and metabolic factors on WM development (Ball et al., 2010; Barnett et al., 2018; Pogribna et al., 2013; Lee et al., 2014; Boardman & Counsell, 2019). Overall, the scarcity of tract-specific effects in the analyses including PMA at scan as covariate suggests that most prematurity-related risk factors primarily act through diffuse influences on global WM maturation, rather than selectively targeting individual tracts.

This motivated a second analytical approach in which variance shared with global WM was considered in order to better isolate tract-specific vulnerability. This strategy was conceptually supported by evidence that a substantial proportion of WM microstructural variance is shared across major tracts in the neonatal brain (Telford et al., 2017). Of note, other diffusion MRI modeling approaches such as fixel-based analysis (Raffelt et al., 2015) have aimed to separate global from tract-specific sources of variance by distinguishing fibre-specific from voxel-averaged signals. Nevertheless, such models could not be applied to our dataset given the image characteristics limited by the clinical context of acquisition (see discussion below).

In our study, a clearer pattern of **tract-specific vulnerability** emerged when global WM was included as a covariate. The sex effect disappeared, consistently with its diffuse and shared impact on global WM and all tracts revealed by the first analyses. Although intracranial volume varied according to sex (estimated adjusted difference of ∼24 mm³ between males and females, p=0.003) and PMA at MRI (estimated adjusted difference of ∼14 mm³ per additional week, p=0.008), the effect of sex on global WM was not driven by differences in brain size, as suggested by additional models including intracranial volume as a covariate (results not shown). On the contrary, the effect of WM abnormality severity (Kidokoro scoring) became more pronounced for projection tracts (CST and STR) when global WM was included as a covariate, indicating that their microstructure is influenced by radiologically visible prematurity-related injury. This suggested that some tracts are intrinsically more vulnerable, likely reflecting differences in developmental timing and anatomical localization, such as the periventricular course of the CST and STR as well as their early maturation compared to other more distal tracts (Kostović et al., 2019).

In contrast to other tracts, the FAT showed effects of GA, SGA at birth and invasive ventilation but in unexpected directions, with higher FA and/or lower MD in infants presenting higher clinical risk after adjustment for global WM microstructure. One potential but superficial interpretation could be a paradoxical “over maturation” of FAT in these vulnerable infants, but that would be without considering the effect of the WM adjustment. These observed patterns might rather reflect that, at a comparable global microstructural level, FAT was relatively preserved (higher FA and/or lower MD) compared with the globally affected WM. This would suggest that diffuse insults may have affected this tract less strongly (Tu et al., 2008), but this interpretation nevertheless remains tentative. To examine whether these observations could reflect systematic differences in tract reconstruction between groups, voxel-wise bootstrap analyses were performed on the tract masks of the left and right FAT separately. These indicated no significant differences in FAT localization between G1 and G2-G3 infants, nor between infants with and without invasive ventilation, nor between SGA and non-SGA infants (results not shown), suggesting no bias in FAT reconstruction. If the observed pattern for FAT reflects a biological difference, one possible but unconfirmed explanation may involve the delayed maturation and greater plasticity of frontal associative pathways. Importantly, the FAT has only recently been recognized as a core intra-hemispheric associative component of the frontal-motor network, present in both hemispheres and connecting the inferior frontal gyri with medial frontal motor areas including the supplementary motor area (Catani et al., 2012). Although recent fetal diffusion MRI work has begun to characterize its developmental trajectory (Calixto et al., 2025), FAT microstructure remains poorly characterized in newborns and preterm infants. Current knowledge therefore derives largely from studies in adults and older children, where FAT is implicated in speech initiation and production, motor planning and executive functions (Catani et al., 2013; Dick & Tremblay, 2012). The biological significance of the observed associations therefore remains uncertain and should be confirmed in independent cohorts.

On the other hand, the absence of tract-specific effects in commissural tracts (FMI, MCP) after adjustment for global WM suggested that their microstructural alterations are captured by the diffuse component of WM dysmaturation. This is in line with previous diffusion MRI studies showing that corpus callosum is affected in very preterm infants when compared with term-born controls, but predominantly as part of a global pattern of WM alterations rather than as a specifically vulnerable tract (Thompson et al., 2012). Regarding cerebellum, previous studies in very preterm infants have primarily reported alterations in cerebellar development and cerebello–cortical structural connectivity using global or network-level approaches (Limperopoulos et al., 2005; Limperopoulos et al., 2007; van Kooij et al., 2012; Jeong et al., 2016) but, to our knowledge, MCP has not yet been examined as an individual tract using diffusion MRI in this population. Taken together, the lack of tract-specific effects in the present study might reflect either reduced sensitivity or greater biological resilience of large tracts to subtle microstructural alterations.

Besides, an important aspect that we did not study is the differential development of tracts in the left and right hemispheres which may lead to **inter-hemispheric asymmetries**. To consider tract-level metrics, DTI parameters were averaged within each tract and, for bilateral tracts, across hemispheres, in order to increase signal-to-noise ratio, reduce the influence of local misregistration and facilitate comparisons across infants in this clinically complex cohort, but at the cost of reduced sensitivity to hemispheric asymmetries. Asymmetries are known to emerge early in development, with diffusion MRI studies reporting lateralization of major WM tracts, including the CST, even in infants without overt brain lesions (Dubois et al., 2008). Although systematic neonatal data remain limited and consistent patterns across healthy and clinical populations are still under investigation (e.g., some tracts show structural asymmetries in healthy preterm neonates while others do not (Liu et al., 2010)), emerging evidence also indicates that structural asymmetries evolve dynamically during early infancy rather than following a fixed pattern (Liu et al., 2021). Preserving hemispheric information would therefore provide important insight into how prematurity-related risk factors might modulate the development of structural lateralization (Hervé et al., 2013; Ocklenburg et al., 2015). Furthermore, this might be impacted by the presence and location of early brain lesions and considering the **brain lesion laterality** would be informative to get a comprehensive description of asymmetries development. Nevertheless, in this study, the variability across infants was too important in terms of lesion characteristics to consider these questions. Given this substantial inter-individual variability, we considered averages over left and right sensorimotor tracts to facilitate reproducibility and comparisons across infants when examining associations with prematurity-related risk factors. This allowed us to establish a dedicated framework that can be extended in the future to the fine-grained analysis of inter- hemispheric asymmetries.

### MRI markers of the developing WM microstructure

This study relied on diffusion MRI to reveal **tract-level microstructural signatures** in the developing brain. This approach provides *in vivo* sensitivity to the microstructural organization of WM, capturing differences related to axonal density, fiber coherence and myelination (Dubois et al., 2006; Ouyang et al., 2019). The distinct microstructural profiles that we observed across the studied sensorimotor tracts (particularly CST, STR and FAT) in extremely-to-very preterm infants at TEA confirmed the differential maturation patterns across tracts in line with established WM myelination progression (Dubois et al., 2014) and broader developmental trajectories (Lebel et al., 2018; Kimpton et al., 2021). In the absence of an adult or a full-term control group, direct comparisons of maturation across tracts were not possible. This should be kept in mind, as WM tracts show distinct and persistent microstructural properties in adulthood (Lebel et al., 2012). Accordingly, between-tract differences observed at TEA should be interpreted as relative patterns within the preterm cohort rather than as differential maturational delay relative to an adult or a full-term reference.

Beyond normative maturational variability, diffusion metrics reflect subtle deviations or microstructural alterations associated with prematurity or perinatal risk factors (Boardman et Counsell, 2019). In our cohort, tracts such as the CST, STR and FAT showed patterns suggestive of altered maturation and could serve as candidate markers of early WM vulnerability. Relatedly, the absence of a full-term control group prevented us to determine whether the observed microstructural patterns reflected prematurity-related alterations or fall within the normal range of neonatal variability (Bartha et al., 2007). Consequently, any inferences about alterations should be interpreted in the context of relative differences within the preterm sample, rather than as absolute deviations from a normative baseline.

Within this pathological neurodevelopmental context, microstructural indices were derived from a single-tensor DTI fit. A lower FA or higher values of diffusivities (MD, RD and AD) across this at-risk population likely reflect a combination of delays and/or disturbances affecting oligodendrocyte maturation and axonal growth and thus altering the course of WM myelination and global maturation (Volpe et al., 2011; French et al., 2009; Panfoli et al., 2018). It would have been interesting to quantify other MRI markers (e.g., quantitative T1) to better distinguish between these processes, but the limited acquisition time prevented us from doing so. Interestingly, our post-processing methodology allowed us to explore previously understudied tracts (e.g., FAT), complementing the characterization of some more commonly investigated WM tracts in preterm populations (e.g., CST; Kaur et al., 2014).

We demonstrated that clinically meaningful microstructural information can be extracted in sensorimotor tracts in a very high proportion of extremely-to-very preterm infants, despite the substantial **technical and anatomical challenges inherent to neonatal diffusion MRI and tractography** (Dubois et al., 2021). These findings support the feasibility of tract-level comparisons across infants within the present cohort, although formal test-retest reliability and external reproducibility were not assessed. In neonates, acquisitions are generally performed during natural sleep, under continuous clinical monitoring, and are inherently constrained by spontaneous movements and limited tolerance to prolonged scanning, typically restricting total acquisition time to approximately 45 minutes. In this context, diffusion MRI was deliberately limited to a single-shell acquisition, which remains the standard approach in clinical neonatal MRI. While widely used and clinically feasible, DTI models the water diffusion as a single compartment and cannot resolve crossing, fanning or branching fibres, which are abundant in the developing WM (Wheeler-Kingshott et al., 2009; Vos et al., 2011a). Consequently, FA and diffusivity metrics reflect composite signals influenced by multiple microstructural features, including axonal density, myelination, neurite dispersion and extracellular water, limiting mechanistic specificity (Niu et al., 2025). In the present study, no clear differences in sensitivity were observed between the DTI-derived indices, suggesting that these metrics overlap in terms of the aspects of WM microstructure they capture. Future studies could therefore benefit from including more specific diffusion models and indices (e.g. multi-compartment with multi-shell approaches) to better describe the WM microstructure. Nevertheless, it should be noted that the diffusion tensor model was only used to quantify microstructural properties, but not to perform tractography, where a ball-and-stick model with up to two fiber populations was used (Behrens et al., 2007). This approach partially accounts for crossings between two dominant fiber orientations but does not fully represent more complex configurations involving more than two fiber populations.

Beyond acquisition constraints, the infant brain is characterized by rapid developmental changes, reduced tissue contrast and high inter-individual variability (Dubois et al., 2021), which complicate image registration and tractography reconstruction with pipelines developed for adults or older children (Bastiani et al., 2019). Constraints are amplified in extremely-to-very preterm infants, particularly in the presence of brain lesions, ventriculomegaly or atypical anatomy which introduce local distortions and rendered standard registration pipelines inadequate. Rather than excluding these infants, as is often required in conventional diffusion MRI pipelines (e.g. Parikh et al., 2021), we adapted and refined existing tools to include the infants and systematically analyze all datasets. Spatial standardization for tractography seed definition required the use of a neonatal template (Warrington et al., 2022), but carried a risk of local deformation errors, particularly in case of atypical anatomy. Manually designed masks were applied during registration and tract segmentation to constrain alignment and reduce mislocalisation of regions affected by distortions, although this approach introduced operator dependence and reduced automaticity. Importantly, we did not expect these methodological constraints to affect all tracts equally. Compact and coherently oriented pathways such as the CST or STR could have been more reliably reconstructed, whereas tracts located in regions of high fiber complexity, including associative pathways such as the FAT, were more susceptible to geometry-related biases that may complicate biological interpretation (Vos et al., 2011b). Recognizing these tract-dependent specificities is essential for further interpretation of the findings. Thanks to all these adaptations, this study showed that a carefully adapted tractography methodology can be successfully applied to extract reliable tract-level metrics capturing WM microstructure in a clinically vulnerable and anatomically heterogeneous neonatal population that remains largely underrepresented in diffusion MRI studies.

### Clinical considerations and perspectives

The characteristics of this clinical cohort as well as analytical choices might have influenced the study findings and interpretation, with potential implications for its applicability for early risk stratification and longitudinal prognosis of preterm infants. As a first consideration, it should be noted that the absence of a control group of full-term newborns limited the interpretation of the WM microstructural profiles: these could not be viewed as absolute deviations from normative development but as relative differences within the clinical cohort of preterm infants. This limitation prevented us from determining the extent to which the observed profiles were specifically related to prematurity and deviant from typical development. Nevertheless, it did not prevent us from analyzing the inter-individual variability within the preterm cohort and examining associations with clinical risk factors, which was the primary objective of the study.

Besides, the **relatively modest sample size and substantial inter-individual clinical variability of this cohort** may have limited the detection of some subtle tract-specific associations. We restricted the number of perinatal and clinical variables included in the statistical models, in accordance with a previous volumetric study conducted in the same cohort (Elbaz et al., 2025), and based on the literature (Barnett et al., 2018; Boardman et Counsell, 2019) and clinical expertise. Concerning the chosen clinical factors, GA at birth alone did not fully reflect clinical vulnerability in this cohort, and the absence of a monotonic GA effect likely relied on the clinical inter-individual heterogeneity of infants classified as lower risk (G2) compared with others (G1 and G3). This suggested that GA-based stratifications are relatively limited and reinforced the importance of integrating postnatal clinical variables and MRI markers when interpreting early WM development. Also, perinatal adverse clinical factors and neonatal morbidities (e.g., sepsis, BPD, invasive mechanical ventilation, prolonged parenteral nutrition) were expected to be associated with altered WM microstructure at TEA. Yet, the number of infants per group might have been too limited to highlight some effects, except the predominant ones, such as the association between the severity of WM abnormality and CST or STR microstructure, consistently with prior evidence linking neonatal insults to disrupted oligodendrocyte maturation, axonal development, and myelination (Back, 2017; Shah et al., 2008). Altogether our findings support the notion that both prenatal factors and postnatal adverse exposures (e.g., parenteral nutrition, invasive mechanical ventilation) contribute to tract-specific vulnerability, in line with previous DTI studies in very preterm infants (Ball et al., 2010; Pogribna et al., 2013; Lee et al., 2014). Optimization of postnatal care may then mitigate tract-level vulnerability (Shah et al., 2008; Shin et al., 2016), even though the presence of these risk factors may be indicative of more global pathologies rather than exposures on which one can act.

Regarding **analytical considerations**, two complementary modelling strategies were implemented: adjustment for PMA at MRI to control for general age-related effects, and adjustment for global WM microstructure to isolate tract-specific associations independent of diffuse WM (dys)maturation. Besides, correction for multiple comparisons was performed to reduce the risk of false-positive findings but it likely decreased sensitivity to small effects. Accordingly, non-significant trends were reported with caution, as they may still reflect biologically meaningful patterns. A conservative analytical strategy was also considered not to keep outlier observations: for tract-metric combinations that did not meet the assumptions of the linear model, Cook’s distance was used to identify and exclude observations exerting undue influence on model estimates, thereby reducing the sample size only for the affected analyses.

Regarding possible **clinical implications** for this study, the CST, STR, and FAT emerged as early candidate markers of WM vulnerability in extremely-to-very preterm infants at TEA because of their residual tract-specific alterations, detectable after accounting for global WM. This suggests that microstructural deviations related to some clinical risk factors can be identified early in life, potentially preceding overt functional impairment. Although the number of infants with moderate or severe Kidokoro scores was limited, integration of conventional MRI scoring with diffusion-based tract markers may improve early risk stratification by capturing global WM injury severity. Finally, MRI examination at TEA for this preterm cohort was part of a still ongoing **multimodal follow-up** for some infants, including MRI at 2 months of corrected age, high-density EEG, and detailed behavioral assessments up to 42 months of corrected age. Longitudinal data are essential to distinguish transient maturational delays from persistent neurodevelopmental alterations and to establish the prognostic value of early WM markers. Early identification of infants at risk for motor, cognitive, or broader neurodevelopmental impairments may ultimately enable more personalized monitoring and timely intervention during a period of intense neuroplasticity.

## Conclusion

This study illustrates the potential of diffusion MRI to capture early inter-individual variability of WM development at TEA in extremely-to-very preterm infants by considering different clinical risk factors. Beyond the global WM dysmaturation related to premature birth, the identification of tract-specific microstructural profiles within the sensorimotor network suggests that early maturation is shaped by both shared and pathway-dependent processes, reflecting the complex interaction between biological maturation and perinatal exposures. Placing these findings within a multimodal and longitudinal framework will be crucial to refine their biological interpretation and clinical relevance, to improve early phenotyping of WM vulnerability and support more individualized care approaches.

## Data availability

Derived data generated for this study will be shared on reasonable request to the corresponding author.

## Author contributions

Conceptualization: LD, JD, MA, VB, MBR

Data curation: LD, NE, CG, JD, MA, MEB, AGC

Formal analysis: LD, YL, JD, NE, AGC

Investigation: LD, NE, CG, JD, PA, MA, MEB, VB, AH

Methodology: LD, YL, JD, SN, JD, MA, VB

Project administration: JD, MA, VB, LHP, MBR, CC

Resources: YL

Supervision: JD, MBR, MA, VB

Validation: Ethic Committee of research in medical imaging, of the CERF (Collège des Enseignants en Radiologie de France), *Comité de Protection des Personnes* (CPP) Ile de France 3

Visualization: LD

Writing – original draft: LD

Writing – review and editing: LD, JD, YL, MA, VB, CC, LHP, MBR, SN

All authors approved the final manuscript as submitted and agree to be accountable for all aspects of the work.

## Ethics approval

The study was approved by the Ethic Committee of research in medical imaging, of the CERF (Collège des Enseignants en Radiologie de France) and the *Comité de Protection des Personnes* (CPP) Ile de France 3 (protocol DEVine, CEA 100 054).

## Consent statement

This study was conducted following the principle of non-opposition for the use of clinical and imaging data for research purposes.

## Acknowledgments

The authors would like to thank clinical teams at Robert-Debré Hospital (in particular M. Riche, M. Toquer, R. Bonicel and radiographers) and at NeuroSpin/UNIACT (in particular G. Mediouni and B. Martins) whose collaboration made it possible to conduct this study in a clinical setting. The authors also thank S.N. Sotiropoulos, S. Warrington and E. Thompson for helpful discussion on the baby-XTRACT tool.

## Abbreviations

AD: Axial Diffusivity
BET: Brain Extraction Tool
BPD: Bronchopulmonary Dysplasia
CSF: Cerebrospinal Fluid
CST: Corticospinal Tract
DEHSI: Diffuse Excessive High Signal Intensity
DTI: Diffusion Tensor Imaging
DWI: Diffusion-Weighted Imaging
EPI: Echo-Planar Imaging
FA: Fractional Anisotropy
FAT: Frontal Aslant Tract
FDR: False Discovery Rate
FMI: Forceps Minor
FOV: Field of View
GA: Gestational Age (at birth)
IVH: Intraventricular Hemorrhage
MD: Mean Diffusivity
MRI: Magnetic Resonance Imaging
mT/m: Milli-Tesla per Metre
NEC: Necrotizing Enterocolitis
PMA: Post-Menstrual Age
PVL: Periventricular Leukomalacia
RD: Radial Diffusivity
ROI: Region Of Interest
SD: Standard Deviation
SGA: Small for Gestational Age (at birth)
T: Tesla
T2w: T2-weighted
TBSS: Tract-Based Spatial Statistics
TE: Echo Time
TEA: Term-Equivalent Age
TR: Repetition Time
TSE: Turbo Spin echo
WM: White Matter

---

**Supplementary Figure 1.**
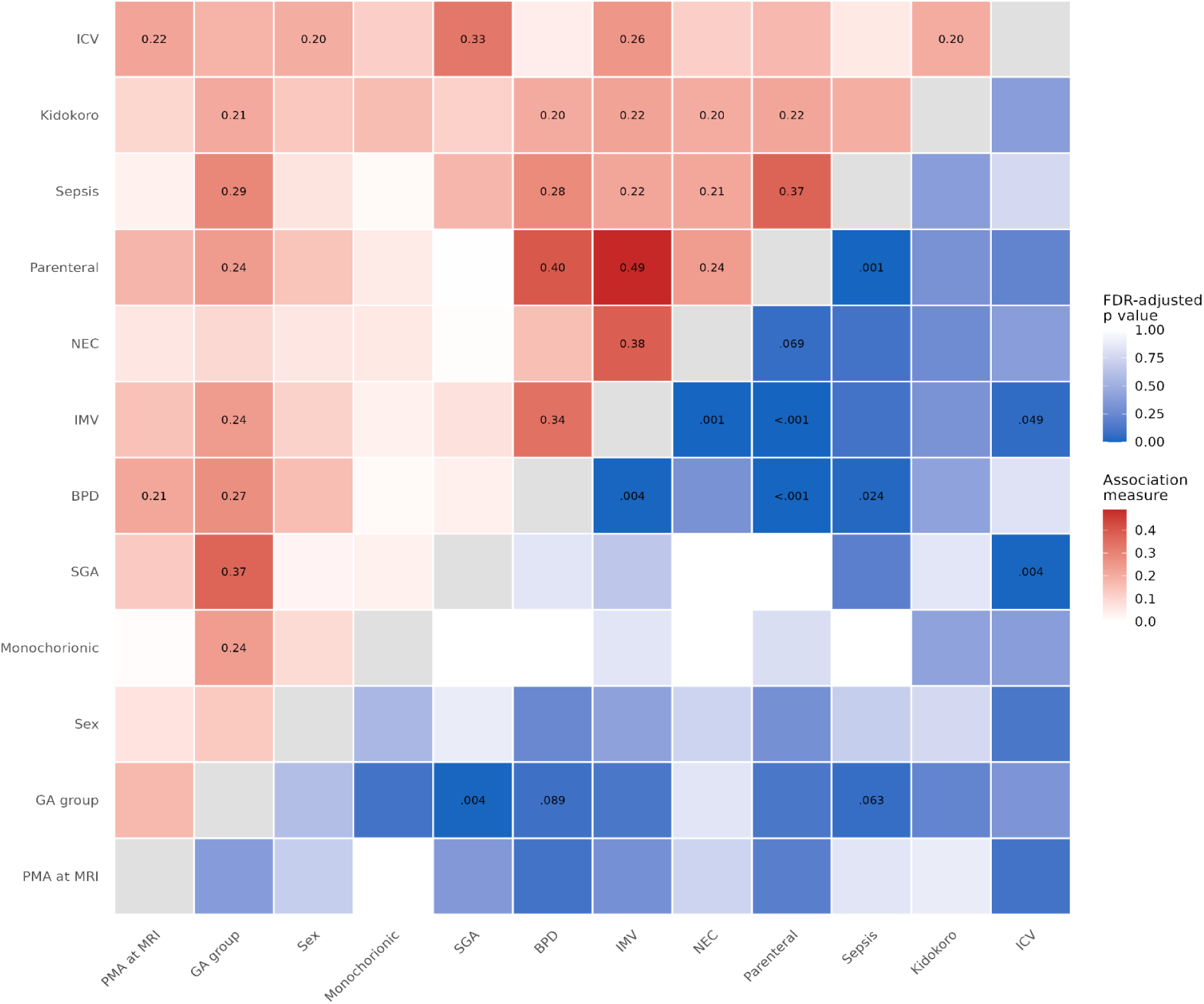
Associations between clinical risk factors and explanatory variables introduced in the statistical models. ***Upper triangle:*** strength of pairwise association, quantified using absolute Pearson’s r, Cramér’s V, or the correlation ratio η, depending on variable types. Values <0.2 are not displayed as they were considered as non-significant for a sample of 111 infants. The shades of red show the association strength. ***Lower triangle:*** FDR-adjusted p values from Pearson correlation tests for continuous-continuous pairs, one-way ANOVA for continuous-categorical pairs, and Pearson’s chi-square or Fisher’s exact tests for categorical-categorical pairs. Fisher’s exact test was used when at least one expected cell count was lower than 5. Only p values ≤ 0.10 are numerically displayed. The shades of blue show the significance strength. ***Abbreviations:*** BPD: bronchopulmonary dysplasia; GA: gestational age (at birth); Kidokoro: Kidokoro severity category of white matter abnormality; Monochorionic: monochorionic twin pregnancy; ICV: intracranial volume; IMV: invasive mechanical ventilation; NEC: necrotizing enterocolitis; Parenteral: prolonged parenteral nutrition; PMA: post-menstrual age; SGA: small for gestational age at birth.

**Supplementary Figure 2.**
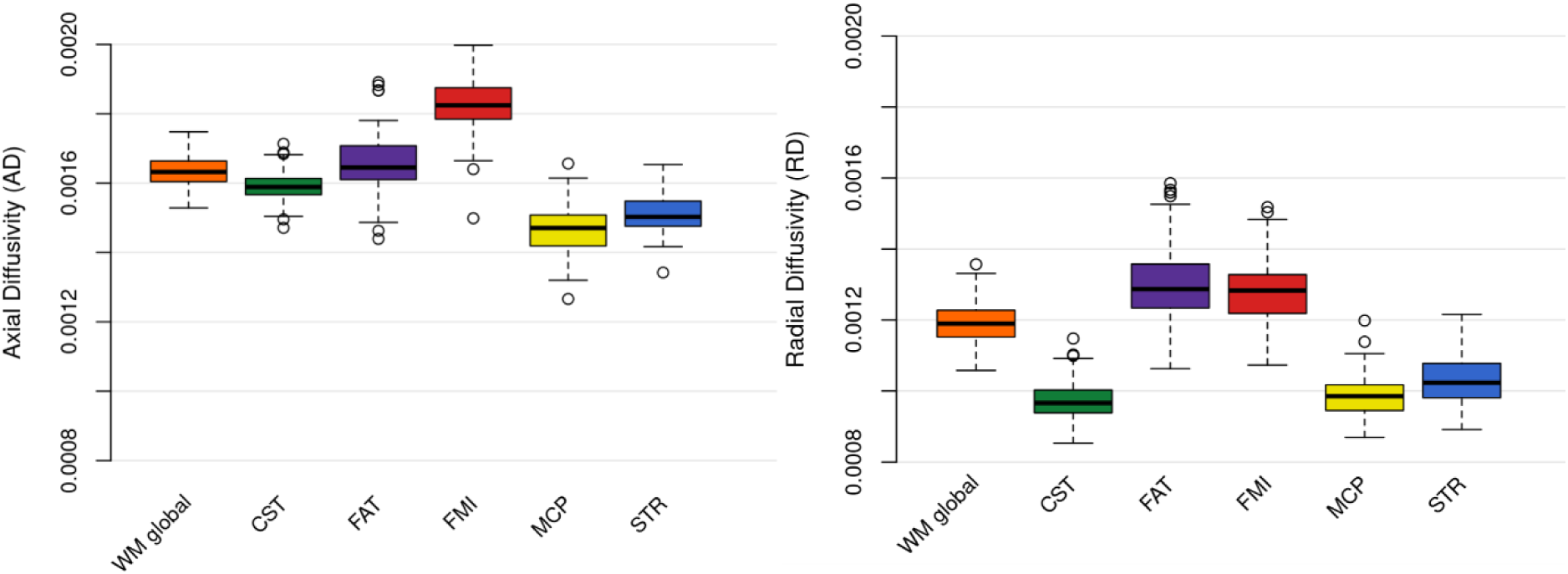
Quantification of the white matter microstructure with axial diffusivity (AD) and radial diffusivity (RD). Box plots of the weighted mean values of AD (left) and RD (right) parameters quantified in the global white matter (42 tracts) and in the 5 tracts of interest, listed in alphabetical order. ***Abbreviations:*** see Table 1 and Figure 4.

**Supplementary Figure 3.**
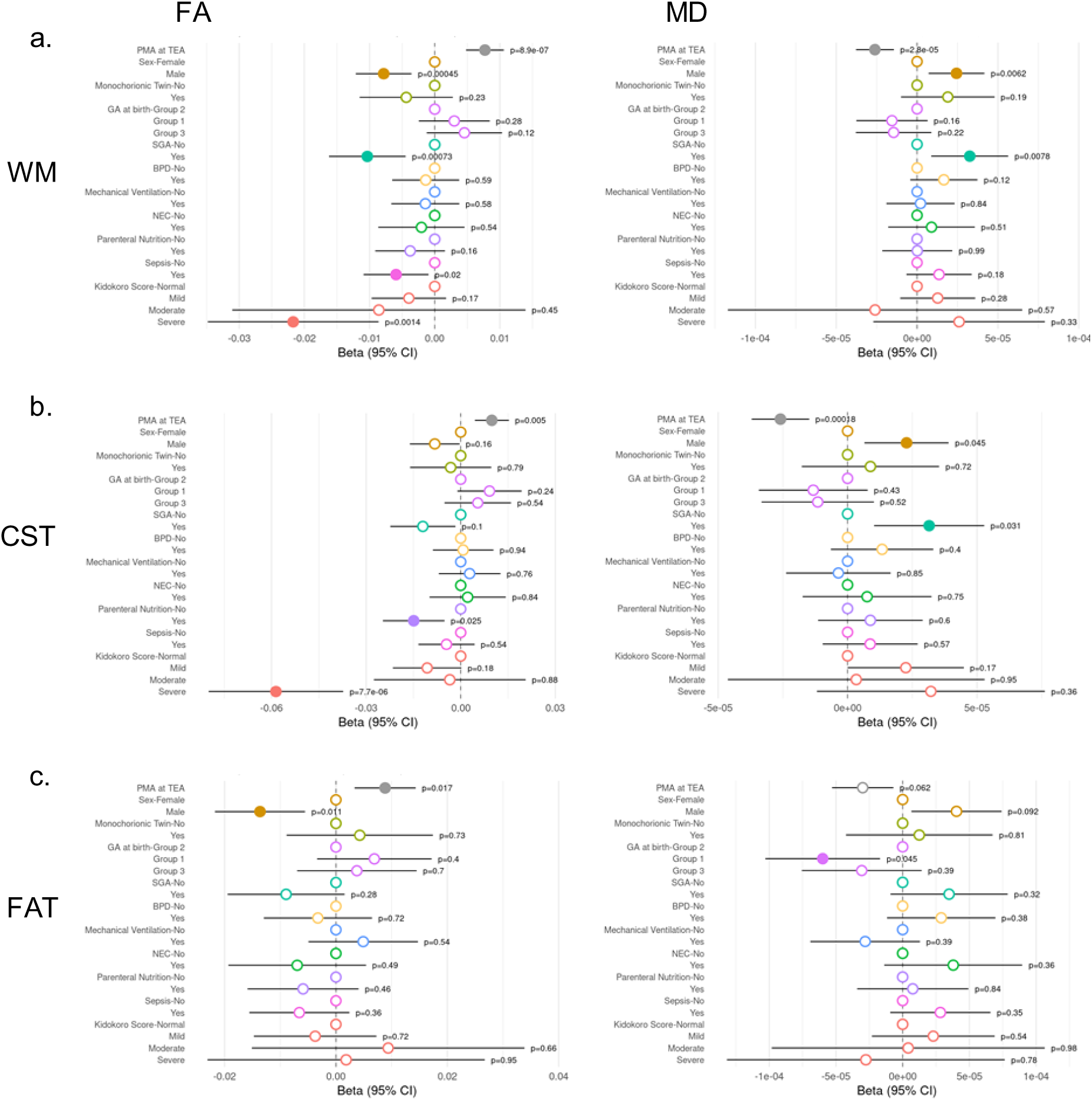
Effects of risk factors and post-menstrual age (PMA) at MRI on fractional anisotropy (FA) and mean diffusivity (MD) over the global white matter (WM) and selected tracts. Illustration of the results of statistical analyses summarized in Table 2.a. for FA (left) and MD (right) quantified over (a) the global WM (N=103), (b) CST (N=110) and (c) FAT (N=110). For binary risk factors, “yes” indicates the presence of the risk factor, whereas “no” indicates its absence and represents the reference category (positioned at 0). p-values were corrected for multiple comparisons over all the global WM and 5 studied tracts, within each model for each DTI metric using FDR approach. Circles location represents the estimated effects, and horizontal lines represent their 95% confidence intervals. Filled circles indicate statistically significant effects after FDR correction at p < 0.05, whereas open circles indicate non-significant effects. ***Abbreviations:*** see Table 1 and Figure 4.

**Supplementary Figure 4.**
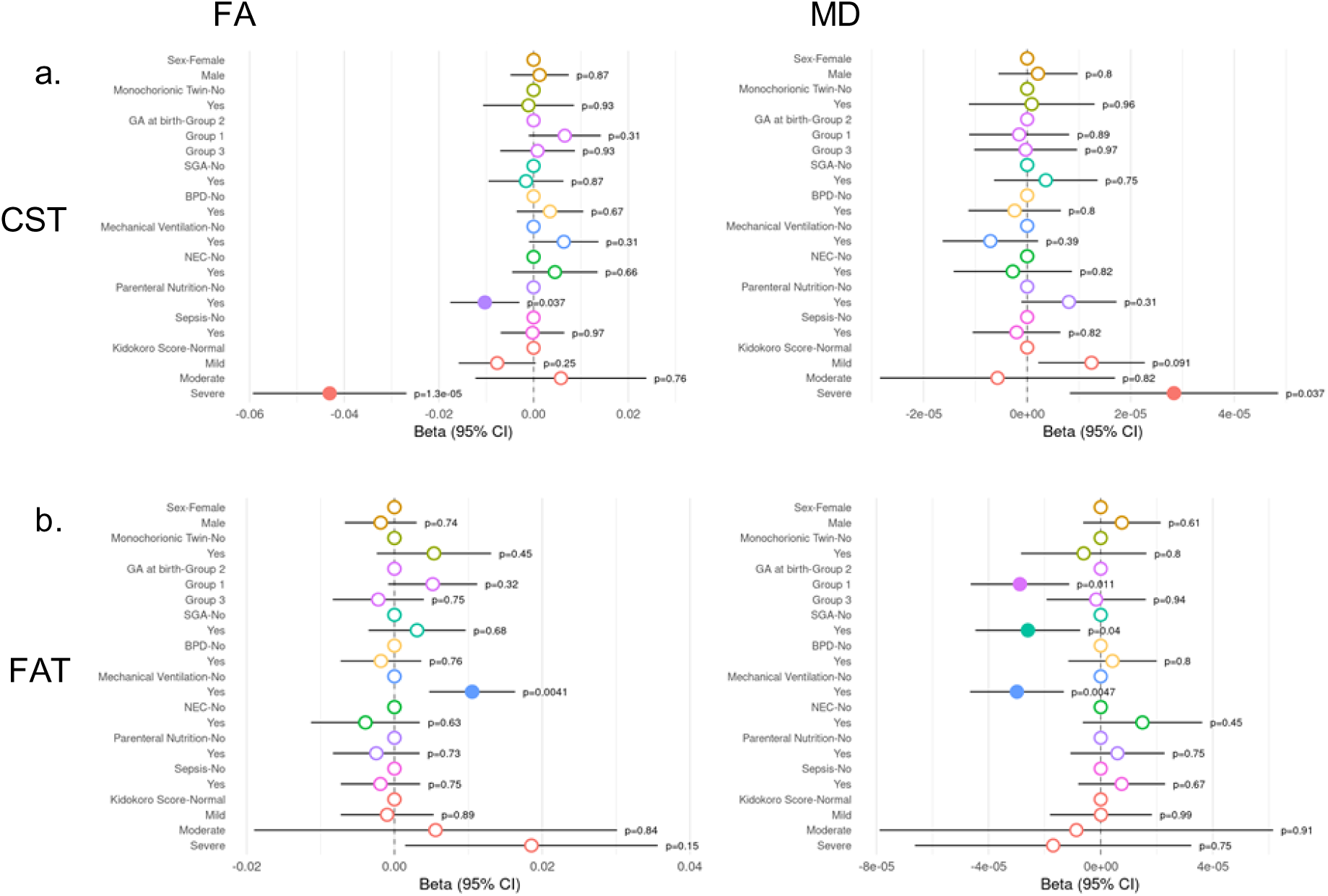
Effects of risk factors and global WM microstructure on FA and MD over the CST and FAT. FA (left) and MD (right) quantified over (a) CST (N=110), and (b) FAT (N=104). For readability, the effect of global WM is not displayed in the forest plots, because of the scale differences between effects. See Supplementary Figure 3 legend for details. ***Abbreviations:*** see Table 1 and Figure 4.

**Supplementary Table 1.**
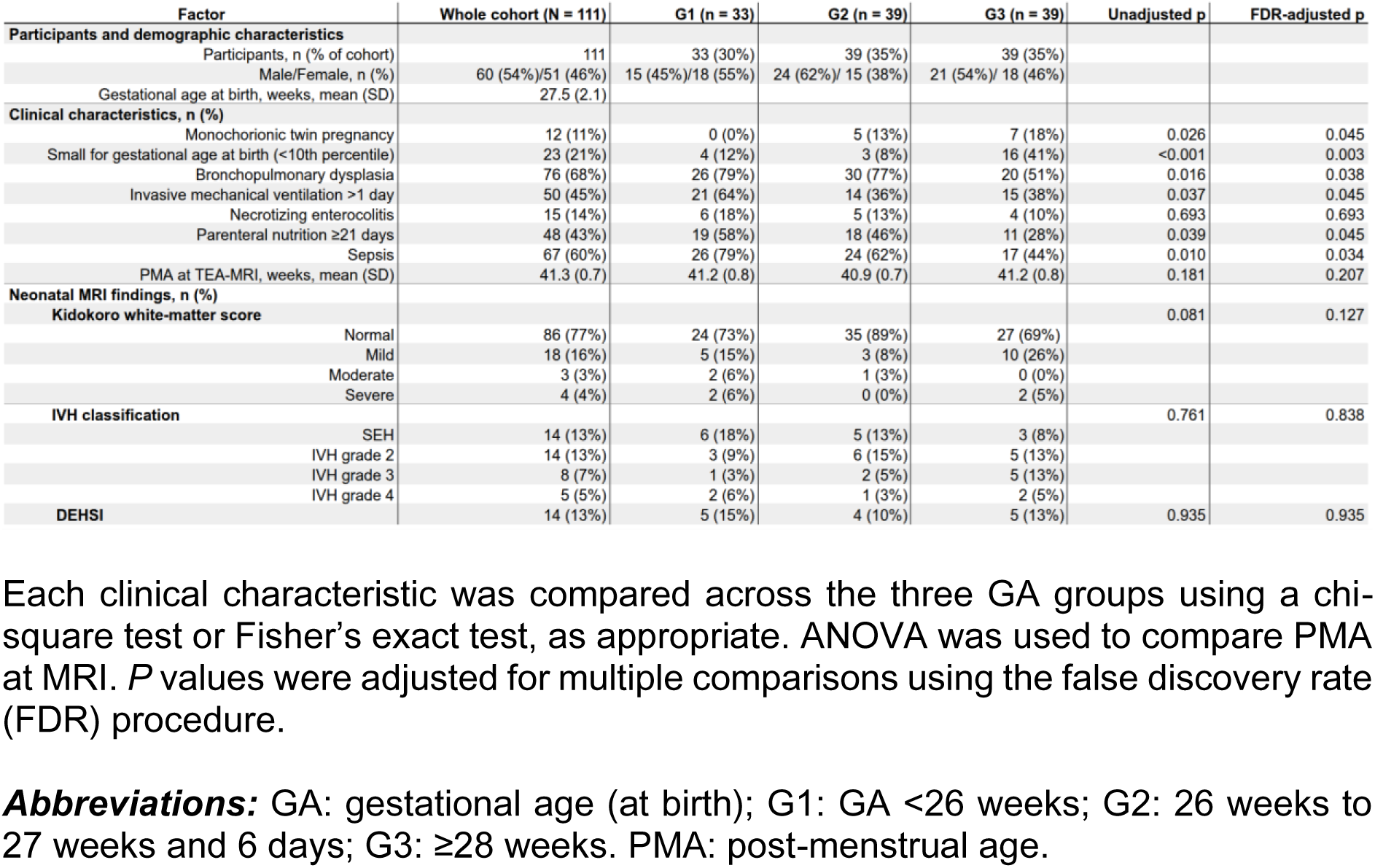
Number and percentage of infants presenting each clinical characteristic within the three groups defined based on GA at birth.

**Supplementary Table 2.** Distribution of unsuccessful tract reconstructions by tract and hemisphere.

| Tract | Right only, n | Left only, n | Both hemispheres, n | Missing in at least one hemisphere, n | Percentage of N = 111 |
| --- | --- | --- | --- | --- | --- |
| AC | 0 | 0 | 12 | 12 | 10.8% |
| AF | 29 | 4 | 5 | 38 | 34.2% |
| AR | 29 | 3 | 5 | 37 | 33.3% |
| ATR | 0 | 1 | 0 | 1 | 0.9% |
| CBD | 5 | 6 | 5 | 16 | 14.4% |
| CBP | 13 | 14 | 7 | 34 | 30.6% |
| CBT | 0 | 0 | 1 | 1 | 0.9% |
| CST | 0 | 0 | 1 | 1 | 0.9% |
| FAT | 1 | 0 | 0 | 1 | 0.9% |
| FMA | 0 | 0 | 0 | 0 | 0.0% |
| FMI | 0 | 0 | 0 | 0 | 0.0% |
| FX | 23 | 15 | 7 | 45 | 40.5% |
| IFO | 0 | 0 | 0 | 0 | 0.0% |
| ILF | 0 | 0 | 1 | 1 | 0.9% |
| MCP | 0 | 0 | 3 | 3 | 2.7% |
| MDLF | 0 | 0 | 0 | 0 | 0.0% |
| OR | 0 | 0 | 0 | 0 | 0.0% |
| SLF1 | 0 | 8 | 1 | 9 | 8.1% |
| SLF2 | 8 | 18 | 17 | 43 | 38.7% |
| SLF3 | 3 | 5 | 2 | 10 | 9.0% |
| STR | 0 | 0 | 0 | 0 | 0.0% |
| UF | 0 | 0 | 1 | 1 | 0.9% |
| VOF | 0 | 0 | 0 | 0 | 0.0% |

The number of infants (n) with unsuccessful tract reconstructions are reported for each tract generated by baby-XTRACT. “Right only” indicates that the tract could not be reconstructed in the right hemisphere but was successfully reconstructed in the left hemisphere. “Left only” indicates the converse. “Both hemispheres” indicates that the tract could not be reconstructed in either hemisphere. “Missing in at least one hemisphere” then corresponds to the sum of these three mutually exclusive categories and represents the number of participants excluded from further analyses. Percentages were calculated relative to the full sample of 111 participants.
**Abbreviations:** AC: anterior commissure; AF: arcuate fasciculus; AR: acoustic radiation; ATR: anterior thalamic radiation; CBD: cingulum subsection: dorsal; CBP: cingulum subsection: peri-genual; CBT: cingulum subsection: temporal; CST: corticospinal tract; FAT: frontal aslant tract; FMA: forceps major; FMI: forceps minor; FX: fornix; IFO: inferior fronto-occipital fasciculus; ILF: inferior longitudinal fasciculus; MCP: middle cerebellar peduncle; MdLF: middle longitudinal fasciculus; OR: optic radiation; SLF1: superior longitudinal fasciculus I; SLF2: superior longitudinal fasciculus II; SLF3: superior longitudinal fasciculus III; STR: superior thalamic radiation; UF: uncinate fasciculus; VOF: vertical occipital fasciculus.

**Supplementary Table 3.** Causes of unsuccessful reconstructions of sensorimotor tracts, involving 4 infants.

| Tract | Side | Number of infants | Likely reason for unsuccessful reconstruction |
| --- | --- | --- | --- |
| FAT | Right | 1 | Cavitary lesion located in the expected FAT region |
| CST | Right and Left | 1 | Registration issue |
| MCP | Non-lateralized | 1 | Registration issue, in the same infant as the CST reconstruction failure |
| MCP | Non-lateralized | 1 | No clear cause identified despite visual quality control |
| MCP | Non-lateralized | 1 | Cerebellar lesion affecting the expected MCP region |

**Abbreviations:** see Supplementary Table 2.

**Supplementary Table 4.**
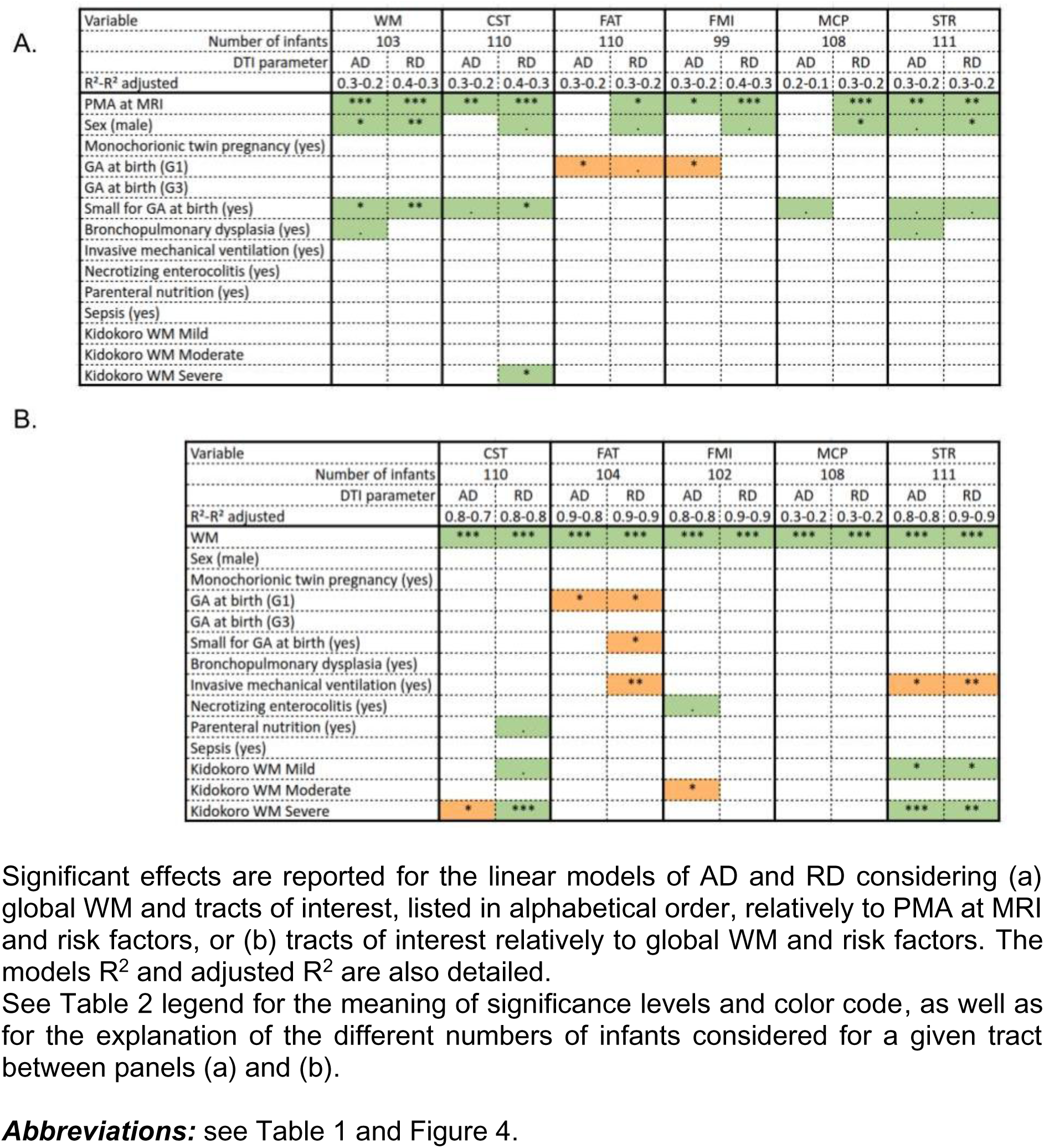
Summary of the results of statistical analyses over the whole cohort for axial diffusivity (AD) and radial diffusivity (RD)

**Supplementary Table 5.**
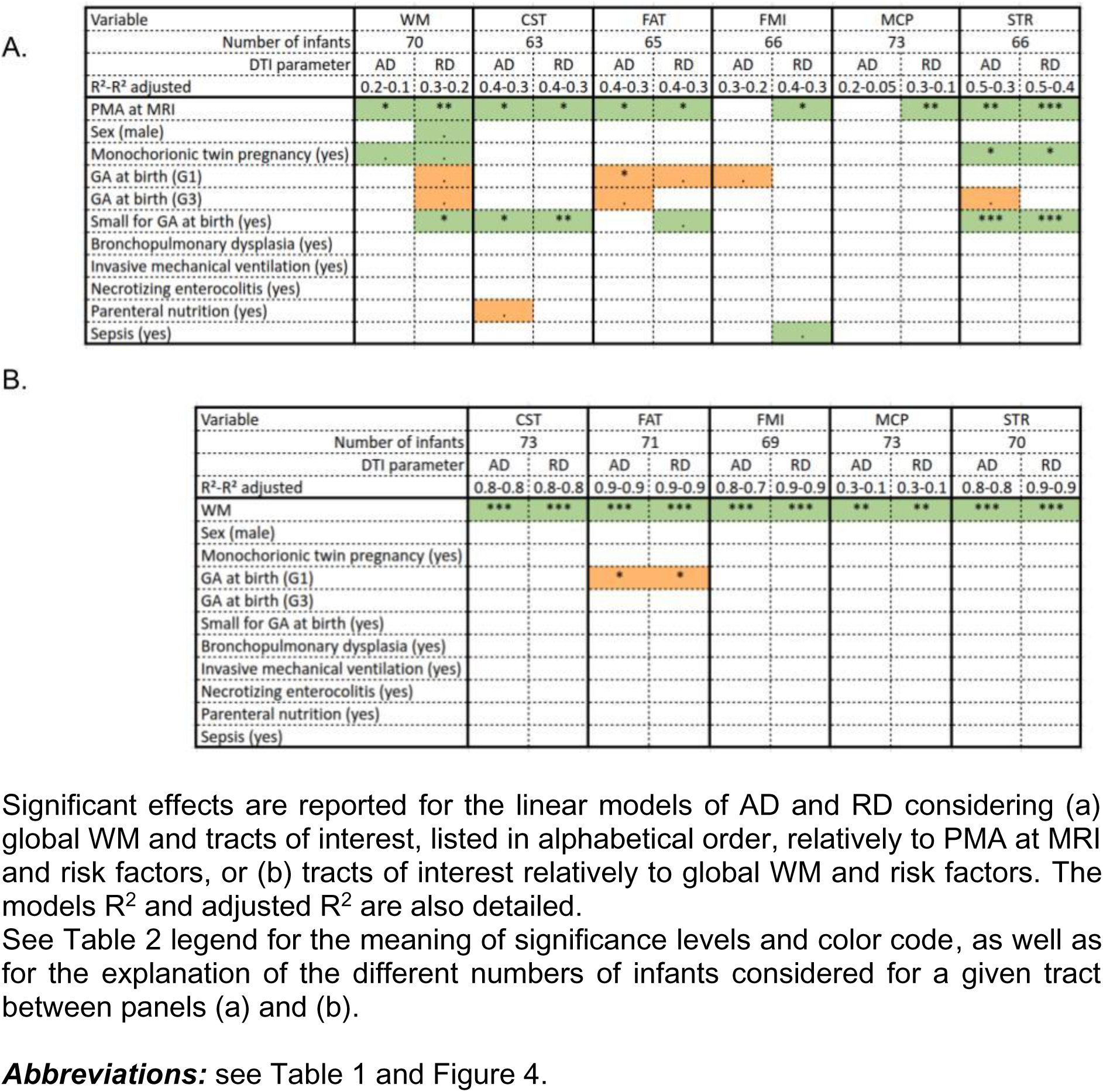
Summary of the results of statistical analyses over the sub-group of infants without macroscopic brain lesions for AD and RD.

